# Altered spike IgG Fc N-linked glycans are associated with hyperinflammatory state in adult COVID and Multisystem Inflammatory Syndrome in Children

**DOI:** 10.1101/2024.07.14.24310381

**Authors:** Jacob D. Sherman, Vinit Karmali, Bhoj Kumar, Trevor W. Simon, Sarah Bechnak, Anusha Panjwani, Caroline R. Ciric, Dongli Wang, Chris Huerta, Brandi Johnson, Evan J. Anderson, Nadine Rouphael, Matthew H. Collins, Christina A. Rostad, Parastoo Azadi, Erin M. Scherer

## Abstract

**Background:** Severe COVID and multisystem inflammatory syndrome (MIS-C) are characterized by excessive inflammatory cytokines/chemokines. In adults, disease severity is associated with SARS-CoV-2-specific IgG Fc afucosylation, which induces pro-inflammatory cytokine secretion from innate immune cells. This study aimed to define spike IgG Fc glycosylation following SARS-CoV-2 infection in adults and children and following SARS-CoV-2 vaccination in adults and the relationships between glycan modifications and cytokine/chemokine levels.

**Methods:** We analyzed longitudinal (n=146) and cross-sectional (n=49) serum/plasma samples from adult and pediatric COVID patients, MIS-C patients, adult vaccinees, and adult and pediatric healthy controls. We developed methods for characterizing bulk and spike IgG Fc glycosylation by capillary electrophoresis (CE) and measured levels of ten inflammatory cytokines/chemokines by multiplexed ELISA.

**Results:** Spike IgG were more afucosylated than bulk IgG during acute adult COVID and MIS-C. We observed an opposite trend following vaccination, but it was not significant. Spike IgG were more galactosylated and sialylated and less bisected than bulk IgG during adult COVID, with similar trends observed during pediatric COVID/MIS-C and following SARS-CoV-2 vaccination. Spike IgG glycosylation changed with time following adult COVID or vaccination. Afucosylated spike IgG exhibited inverse and positive correlations with inflammatory markers in MIS-C and following vaccination, respectively; galactosylated and sialylated spike IgG inversely correlated with pro-inflammatory cytokines in adult COVID and MIS-C; and bisected spike IgG positively correlated with inflammatory cytokines/chemokines in multiple groups.

**Conclusions:** We identified previously undescribed relationships between spike IgG glycan modifications and inflammatory cytokines/chemokines that expand our understanding of IgG glycosylation changes that may impact COVID and MIS-C immunopathology.

## Background

Severe acute respiratory syndrome coronavirus 2 (SARS-CoV-2) infections have a wide range of coronavirus disease (COVID) severities. While many adults infected with SARS-CoV-2 now experience mild COVID, during the early phase of the pandemic, approximately 14% and 5% of adult patients developed severe or critical COVID, respectively[1]. Children and young adults generally present with mild symptoms, but occasionally can have severe disease, and early in the pandemic, 1 out of 3200 pediatric SARS-CoV-2 infections developed into a severe inflammatory disease termed multisystem inflammatory syndrome in children (MIS-C)[2]. MIS-C is considered life threatening[3], though fortunately its incidence seems to be decreasing over time[4, 5]. Severe adult COVID and MIS-C are characterized by excessive inflammation, including elevated inflammatory cytokines and chemokines such as TNFα, IL-6, IL-10, and IL-1β[1, 6–16]. The precise cause of these hyperinflammatory states is incompletely understood[8, 9].

An area of increasing investigation is whether irregular IgG Fc glycosylation is triggering dysregulated inflammation in COVID and MIS-C[17–24]. The IgG Fc region contains a conserved asparagine(N)-linked glycosylation site at N297 on each heavy chain[25]. This glycan is composed of a core seven saccharides containing three mannose and four N-acetylglucosamine (GlcNAc) residues that can be differentially modified with other sugar residues[25] (**Fig. 1**). This diversity of structures has implications for an Fc’s interaction with Fc gamma receptors (FcγRs), which can have downstream signaling effects on an active immune response. For example, it has been demonstrated that afucosylated IgG-Fc have an increased affinity for FcγRIIIa, an activating FcγR found on the surface of monocytes, macrophages, and NK cells, as well as FcγRIIIb, an activating FcγR found on neutrophils[26–30]. The abundance of afucosylated IgG rises in multiple virus infections, including HIV, cytomegalovirus, and dengue[23, 31]. Afucosylated spike IgG1 is also more prevalent in severe than mild cases of COVID[17, 19, 21, 23], with elevated afucosylated IgG1 predicting disease progression[17]. Moreover, afucosylated SARS-CoV-2 spike IgG immune complexes stimulate e*x vivo* human monocytes to secrete inflammatory cytokines[21] and drive inflammatory responses in mouse models expressing human FcγRs[17].

**Figure 1:**
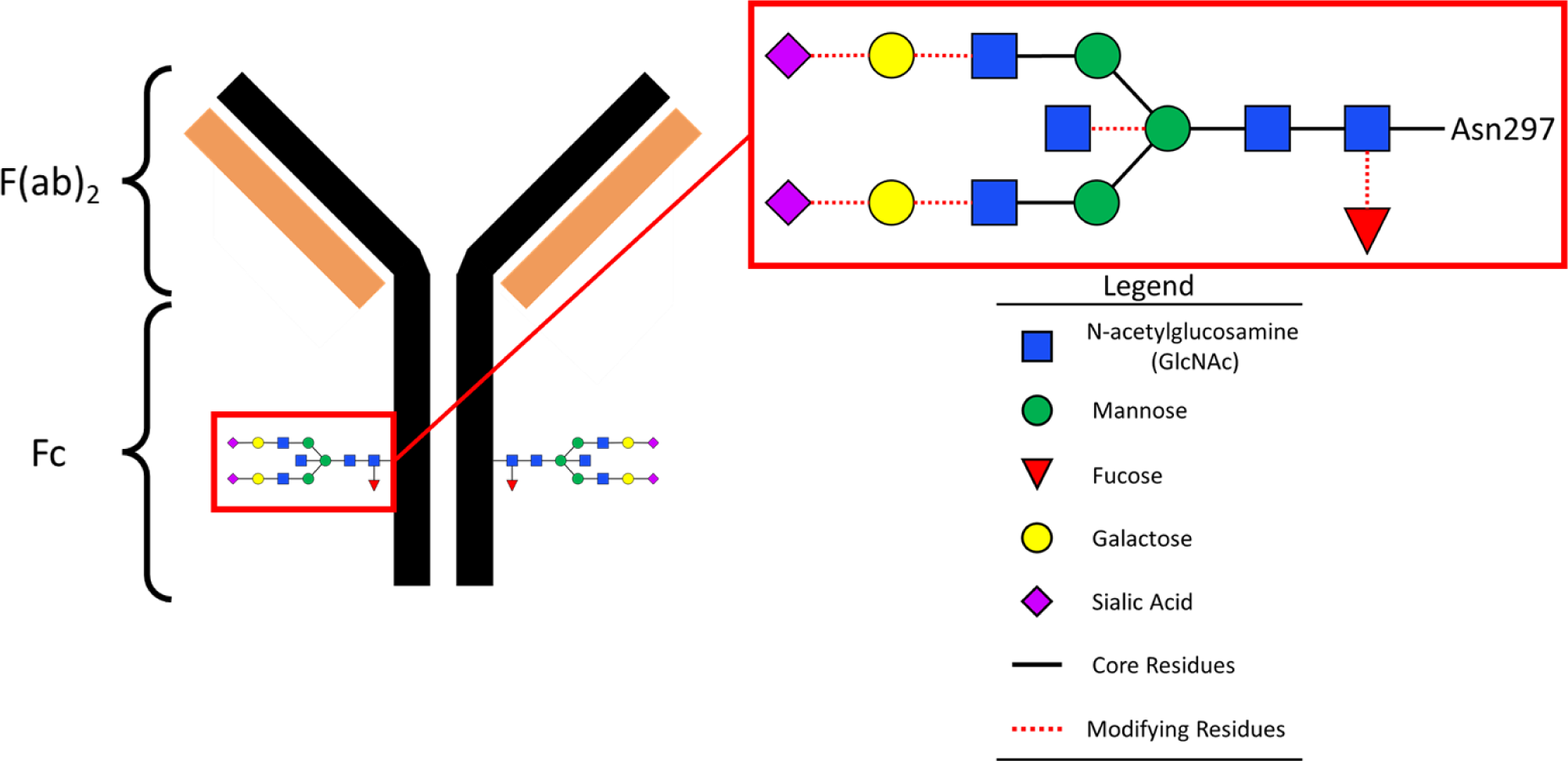
Schematic of IgG structure with conserved N-linked complex glycan at residue 297. IgG is composed of the antigen binding domains, F(ab)2, and constant domain, Fc. The Fc has a highly conserved complex N-glycan at Asn297, which is composed of a core with seven GlcNAc and mannose sugars and additional, variable modifying sugars.

The role of other N297 glycan modifications in inflammation, namely sialylation, galactosylation, and bisection, remains less clear[25]. Studies indicate that Fc sialylation and galactosylation are anti-inflammatory, and that bisecting GlcNAc is positively associated with inflammation[25, 32–34]. However, the findings related to sialylation are discordant among studies and definitive causal relationships between these glycan modifications and changes in inflammation are lacking[25, 35]. Prior work has found SARS-CoV-2 spike IgG more galactosylated and sialylated and less bisected than bulk IgG from the same specimens[19, 23]. These studies have also found bulk and spike IgG are generally less galactosylated, sialylated, and bisected in severe COVID than mild COVID, though these comparisons did not always reach significance[19, 23].

Taken together, these studies suggest afucosylated spike IgG plays a role in inflammation during severe COVID, and we hypothesized this may also extend to MIS-C because of its post-infectious hyperinflammatory profile. When this study was initiated, it was also unknown whether SARS-CoV-2 vaccination led to similar changes in spike IgG glycosylation. The primary objective of this study was to determine whether glycosylation changes are simply byproducts of the antigen specific response and thus also stimulated by vaccination, or if glycosylation changes are unique to MIS-C and COVID and thus potentially driving severe inflammation.

## Materials and Methods

A comprehensive description of materials and methods for this study are included in the Supplementary Information.

### Samples

A breakdown of the study groups is shown in **Supplementary Table 1**. Use of the described secondary research samples for this study was approved by Emory University Institutional Review Board (STUDY00002583).

### Cytokine/Chemokine Multiplex ELISA

Cytokine and chemokine levels were quantified using a custom Mesoscale Discover (MSD) U-plex assay.

### Bulk IgG Isolation, IdeZ Digest, and Fc Enrichment

Heat inactivated serum or plasma samples (200 µL[36]) were desalted with Zeba Spin Desalting plates (ThermoFisher), and bulk IgG purified using Melon Gel Spin plates (ThermoFisher). Twenty-two µL of purified bulk IgG were separated into F(ab)_2_ and Fc fragments by digestion with IdeZ Protease (New England Biolabs). Bulk IgG digests were incubated with Protein G Dynabeads to enrich IgG Fc, and Fc eluted by adding 0.1 M citric acid (pH 3.0) neutralized with 1 M sodium carbonate-bicarbonate buffer (pH 10.8).

### Anti-SARS-CoV-2 Spike IgG Isolation and IdeZ Digest

Anti-SARS-CoV-2 spike IgG enrichment was performed with 50 µL His-tag Dynabeads (Invitrogen 10104D) coated with 12 µg His-tagged recombinant SARS-CoV-2 spike in three rounds. Remaining bulk IgG samples were applied to Dynabead-spike complexes and supernatants transferred to the next set of beads (for 2^nd^ and 3^rd^ enrichments) or discarded (after the 3^rd^ enrichment). Beads from each round of enrichment were pooled for Fc digestion with IdeZ. These IdeZ digests proceeded directly to PNGaseF digest.

### PNGaseF Digest

N-linked deglycosylation of bulk or spike IgG Fc were performed with Rapid PNGaseF (New England Biolabs). After incubation, PNGaseF digests were allowed to dry completely overnight in a biosafety cabinet. Once dried, plates were sealed and stored at 4°C until fluorescent labelling.

### Fluorescent Labelling of IgG Fc Glycans

Dried PNGaseF digests were reconstituted in molecular biology grade water. A solution of 20 mM 8-aminopyrene trisodium salt (APTS) (Sigma-Aldrich) and 3.6 M citric acid (Sigma-Aldrich) was prepared and mixed in equal volume with 200 mM 2-picoline borane (Sigma-Aldrich) in dimethyl sulfoxide (Sigma-Aldrich). This solution was added to each sample and incubated at 37°C for 16 hours, then quenched with 20% ultra-purified water, 80% acetonitrile (Sigma-Aldrich) (v/v). Hydrophilic interaction chromatography based solid phase extraction (HILIC-SPE) was performed to isolate APTS-labelled N-glycans from other reagents as described previously[37]. APTS-labeled samples were stored at -20°C until CE analysis.

### Capillary Electrophoresis

APTS-labeled samples were analyzed using an ABI3130XL or ABI3500XL instrument (Applied Biosystems) with an APTS-labeled N-glycan reference panel (Agilent) run in parallel.

Resulting electropherograms were analyzed using GeneMarker (v3.0.1). Area-under-the-curve (AUC) was quantified for each peak to determine glycoform abundance out of total glycan AUC.

### Mass Spectrometry

An IgG Fc sample (5 µg) was desalted and further reduced and alkylated with 5 mM DTT and 10 mM iodoacetamide. Proteins were digested with a 1:20 ratio of trypsin at 37°C for 18 hours. The resulting peptides were subjected to liquid chromatography with tandem mass spectrometry (LC-MS/MS) on an Orbitrap Eclipse mass spectrometer (ThermoFisher) equipped with 3000 RSLCnano system. Data were acquired in positive ion mode and processed with Byonic software (Protein Metrics; v4.0.12) and searched against the IgG Fc sequence and a catalogue of more than 59 human N-linked glycans. The relative percentages of each glycoform were determined by deconvolution of the LC-MS data at the full MS level, then determining the AUC for each full MH+.

### Statistical Analysis

Statistical analyses were performed in GraphPad Prism (v9.4.1).

## Results

We evaluated cytokine/chemokine levels, bulk IgG Fc glycosylation, and spike IgG Fc glycosylation in sera/plasma specimens from cross-sectional or longitudinal groups of adult healthy controls (n=10), COVID patients (n=109), and mRNA SARS-CoV-2 vaccine study participants (n=37), as well as cross-sectional groups of pediatric healthy controls (n=10) and COVID (n=9) and MIS-C patients (n=20) (**Supplementary Table 1**). The adult COVID group was comprised of patients with severity ranging from moderate (ordinal scale 4, OS4) to severe and critical (OS5 and OS7). COVID patient samples were collected during acute hospitalization for COVID (at enrollment; ≤12 days post-symptom onset) and during convalescence (29 days post-enrollment). Acute samples were first analyzed by ELISA for seropositivity to SARS-CoV-2 spike protein, with 89/109 (82%) considered seropositive by criteria described in the extended methods of the **Supplementary Information**. Seronegative acute samples were omitted from spike IgG Fc glycosylation analysis.

### Cytokine Analysis

We evaluated systemic inflammatory cytokine and chemokine (IL-10, IP-10 (CXCL10), IL-8 (CXCL8), IFNγ, IL-6, MIP1-α (CCL3), IL-1β, MCP-1 (CCL2), MCP-3 (CCL7), and TNFα) levels in all specimens. Adult acute COVID patients exhibited significantly increased cytokine/chemokine levels compared to healthy controls and SARS-CoV-2 mRNA vaccinees for 9/10 of the markers measured, but not IL-1β (**Fig. 2, Fig. S1**). IL-10, IL-6, MIP-1α, MCP-1, MCP-3, and TNFα were significantly elevated in critical COVID (OS7) compared to moderate COVID (**Fig. 2, Fig. S1**). During convalescence, cytokine/chemokine levels generally approached healthy control baseline levels, and this decrease was significant in most cases for adult severe COVID patients who received either remdesivir or placebo, as well as in adult moderate COVID patients who received remdesivir (**Fig. S2, S3**). Among vaccine groups, serum cytokine levels did not significantly change between baseline and 7 days post second vaccine dose, except for a significant increase in IP-10 (**Fig. S4**).

**Figure 2:**
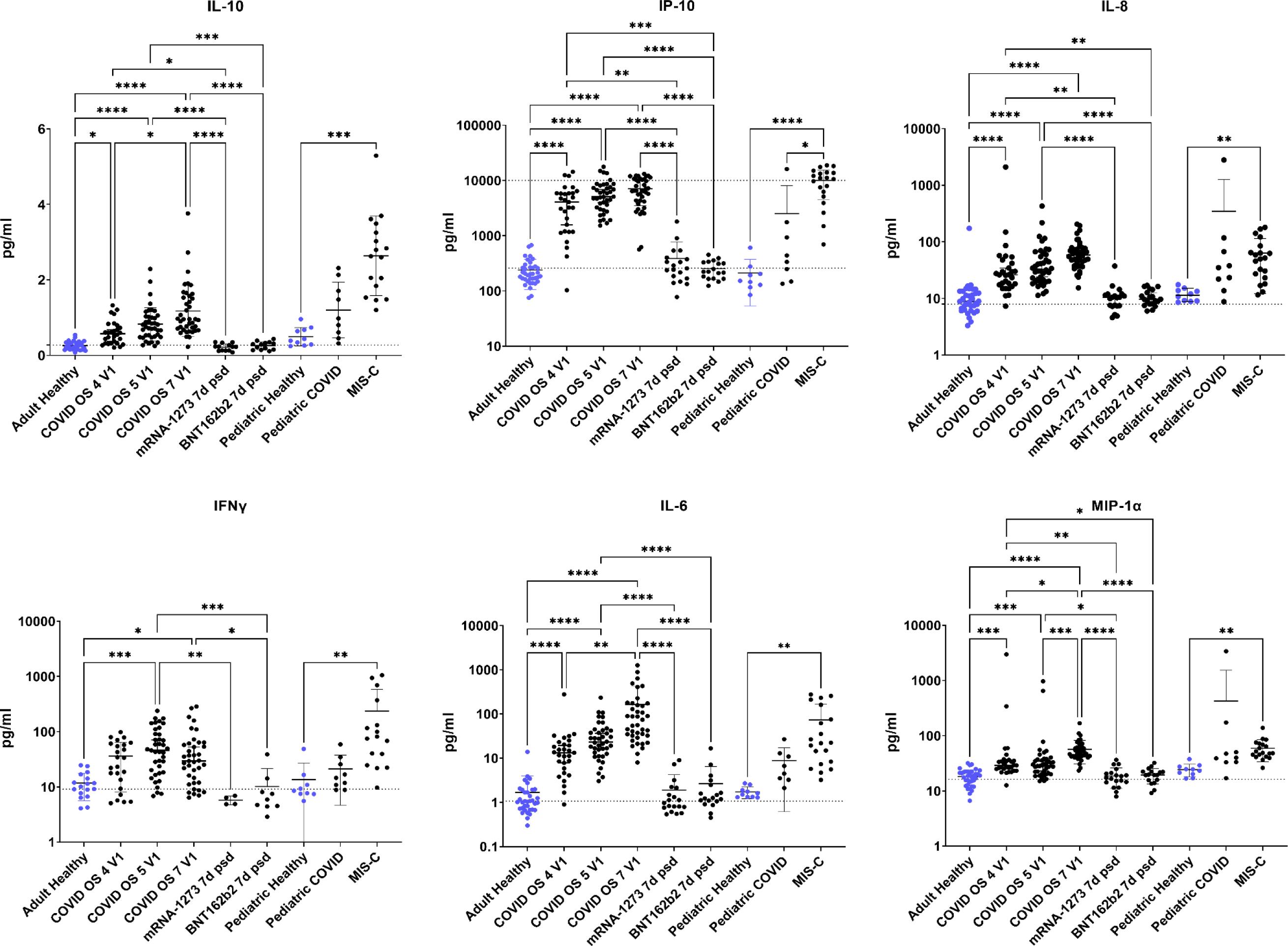
Proinflammatory cytokines and chemokines are significantly elevated in adult COVID patients and MIS-C patients compared to healthy controls. Differences in IL-10, IP-10, IL-8, IFNγ, IL-6, and MIP-1α levels between adult healthy controls, which includes vaccinees at baseline, patients enrolled in ACTT-1 with different COVID severities by ordinal scale (OS) at enrollment (V1), or vaccinees (7 days post-second dose, 7d psd); as well as between pediatric healthy controls or pediatric patients with SARS-CoV-2 or MIS-C. Kruskal-Wallis with Dunn’s test (* p<0.05, ** p<0.01, *** p<0.001, **** p < 0.0001)

Among pediatric study groups, MIS-C patients exhibited significantly increased IL-10, IP-10, IL-8, IFNγ, and MIP-1α compared to healthy controls (**Fig. 2**).

### IgG Fc Glycan Analysis

To evaluate whether IgG Fc glycosylation was altered following SARS-CoV-2 infection or mRNA vaccination, we developed methods for analyzing IgG Fc glycosylation by adapting previously published methods[37–39]. We first purified bulk IgG from serum and then digested it with IdeZ protease to separate the F(ab)_2_ region from the Fc region. We did this to reduce confounding results from glycosylation in the IgG F(ab)_2_ region, which occurs in ∼30% F(ab)_2_ domains[25]. Protein G beads enriched the proportion of Fc domain in the sample but did not remove F(ab)_2_ domain completely (**Fig. S5**). To purify spike IgG for antigen-specific IgG Fc glycan analysis, we treated bulk IgG three times with spike-coated beads. We confirmed spike IgG was depleted from bulk IgG by ELISA (**Fig. 3A**) and that eluted spike IgG bound to spike (**Fig. 3B**). To isolate the Fc domain from spike IgG, we treated IgG bound to spike-coated beads with IdeZ protease, releasing pure Fc and IdeZ with minimal contaminating F(ab)_2_ as confirmed by SDS-PAGE (**Fig. S6**). We then compared the concordance of bulk IgG Fc glycosylation results generated by our CE method with those generated by mass spectrometry (LC-MS/MS) using biological replicate samples, where LC-MS/MS selectively identifies glycans at position N297 (i.e., does not include N-linked glycans in the F(ab)_2_ domain). This comparison showed good general agreement between results obtained by both methods, indicating minimal differences despite F(ab)_2_ contamination in our bulk IgG Fc preparation, though we found a higher proportion of galactosylated glycans by CE (**Table 1**). Independent analyses of replicate bulk IgG Fc samples by CE showed high reproducibility, with a 2-3% standard deviation (**Table S2**).

**Figure 3:**
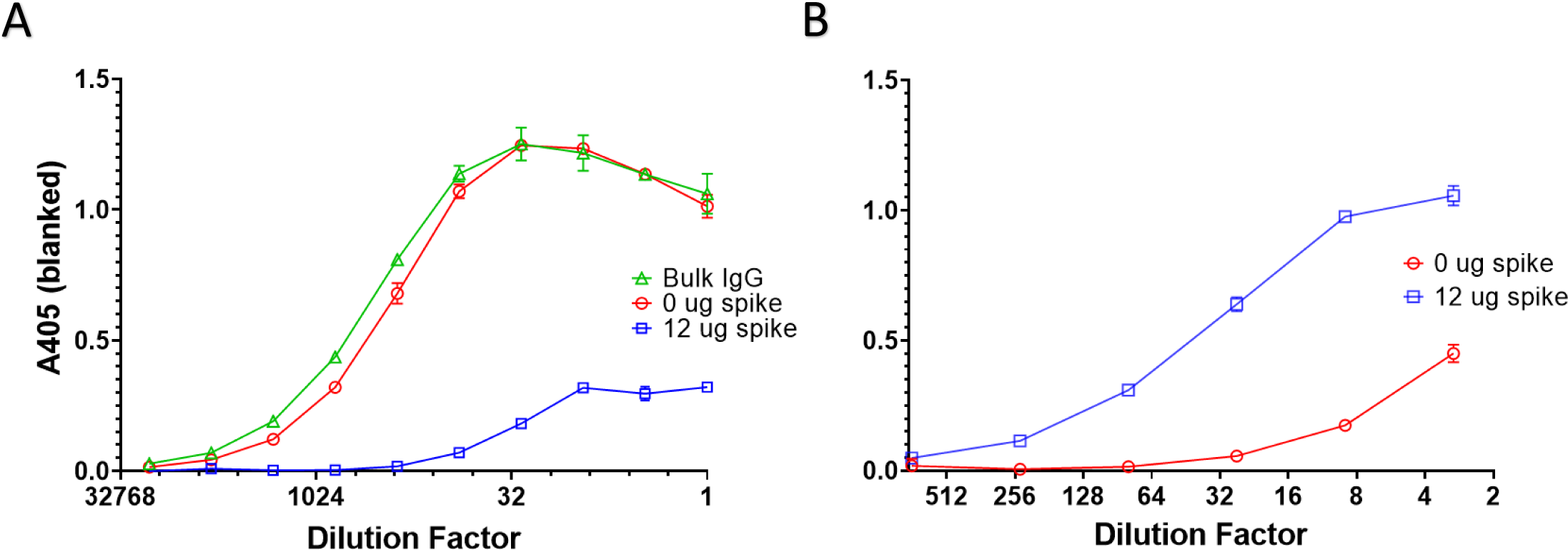
Spike-coated beads capture anti-spike IgG from purified bulk IgG of a seropositive participant. **A)** After treatment three times with His Dynabeads coated with 0 ug (red) or 12 ug (blue) SARS-CoV-2 spike protein, depleted bulk IgG were analyzed for spike binding by ELISA alongside the purified bulk IgG **B)** Measurement of spike binding by post-enrichment IgG eluates from His Dynabeads coated with 0 ug (red) or 12 ug (blue) spike.

**Table 1:**
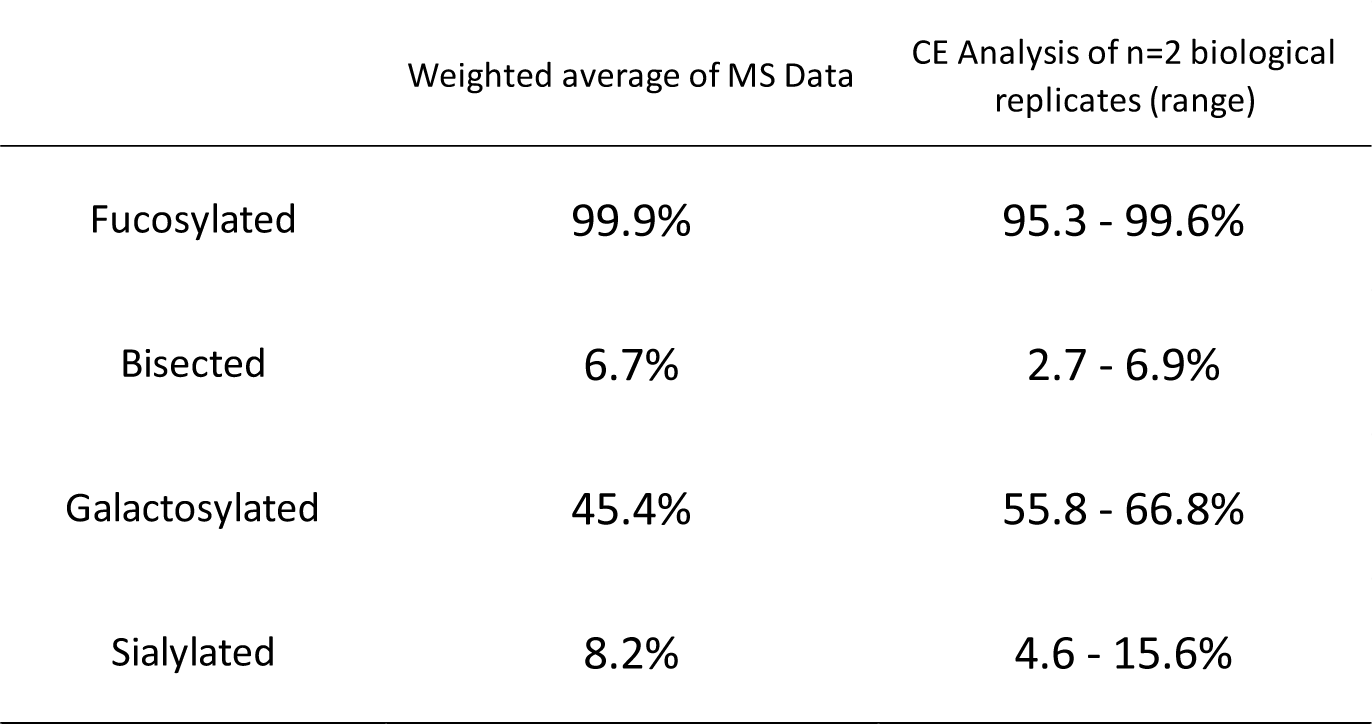
Capillary electrophoresis recapitulates sialylated and bisected bulk IgG Fc glycan abundances found by LC-MS/MS in biological replicate analysis.

|  | Weighted average of MS Data | CE Analysis of n=2 biological replicates (range) |
| --- | --- | --- |
| Fucosylated | 99.9% | 95.3 - 99.6% |
| Bisected | 6.7% | 2.7 - 6.9% |
| Galactosylated | 45.4% | 55.8 - 66.8% |
| Sialylated | 8.2% | 4.6 - 15.6% |

We then analyzed bulk IgG Fc glycans by CE for all samples and spike IgG Fc glycans for seropositive samples. We grouped glycans into four standardized categories: absence of core fucose (afucosylated), presence of terminal galactose (galactosylated), presence of terminal sialic acid (sialylated), or presence of bisecting GlcNAc (bisected). Four glycoforms did not separate adequately by CE, and we observed peaks in samples that were not present in our glycan standards. Thus, we conservatively determined ranges of glycan abundances for each sample, with the minimum limited to only positively identified peaks and maximum including ambiguous peaks. The mean of this range was used for statistical comparisons.

Overall, we found spike IgG were more afucosylated in acute COVID than spike IgG during the peak antibody response to the second SARS-CoV-2 vaccine dose, than bulk IgG from the same groups, or than bulk IgG of healthy controls. Specifically, we observed spike IgG Fc N-linked glycans were significantly more afucosylated in severe OS5 COVID compared to bulk IgG Fc glycans from the same group or adult healthy controls (**Fig. 4**). The proportion of afucosylated spike IgG in moderate COVID was also greater than that observed in BNT162b2 or mRNA-1273 vaccinees. We observed that spike IgG were more afucosylated in moderate or critical (OS7) COVID than bulk IgG from the same groups or adult healthy controls, but these differences were not significant. Spike IgG in MIS-C patients were also more afucosylated than bulk IgG from the same groups or bulk IgG of pediatric healthy controls, but the latter did not reach significance (**Fig 4**.).

**Figure 4:**
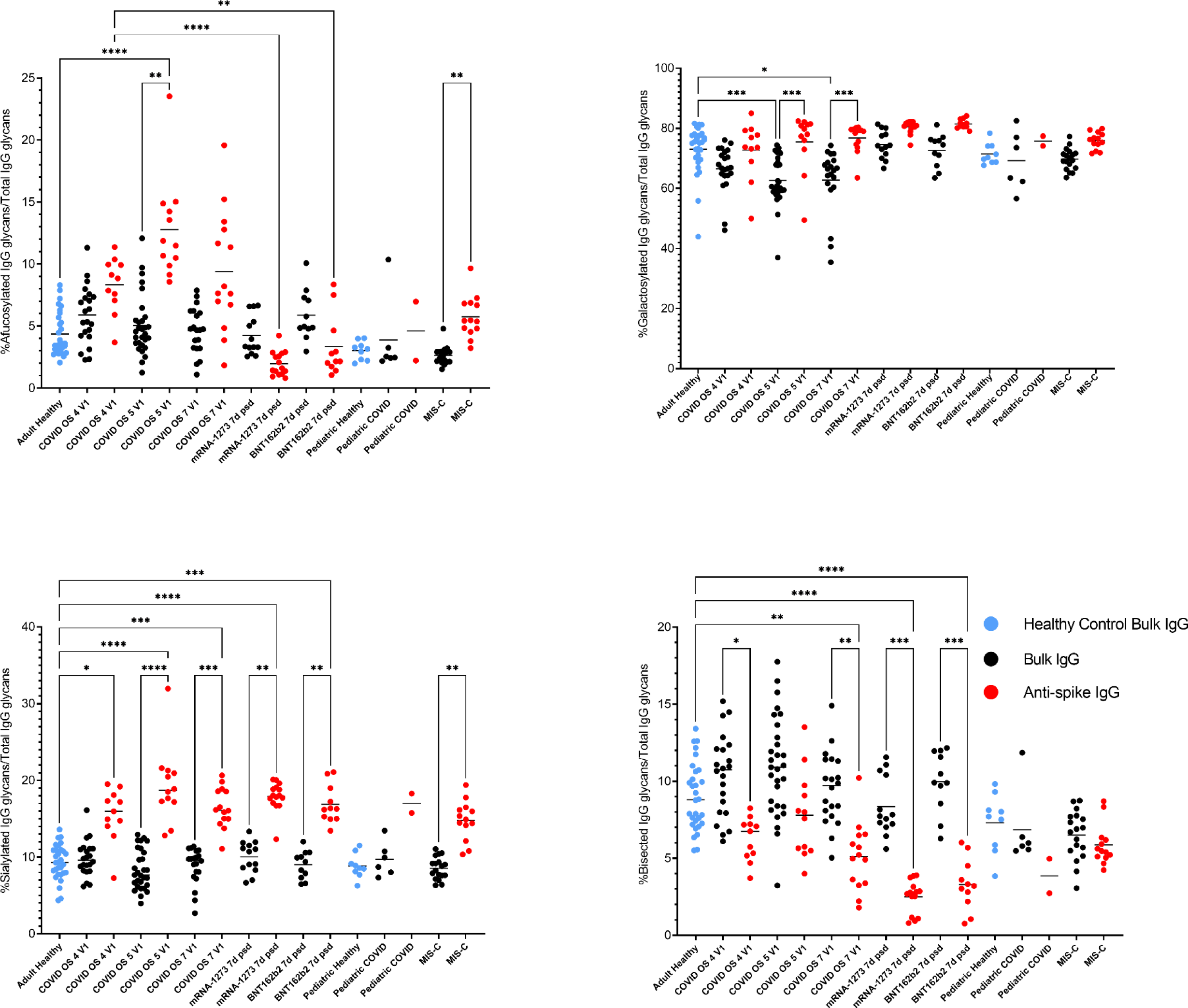
Afucosylated anti-spike IgG are significantly increased in severe COVID and MIS-C over bulk and healthy controls, while anti-spike IgG is globally more sialylated and less bisected than bulk or healthy control IgG. Kruskal-Wallis with Dunn’s test (* p<0.05, ** p<0.01, *** p<0.001, **** p < 0.0001).

In contrast to the opposing trend in spike IgG Fc afucosylation observed during COVID/MIS-C compared to vaccination, we found spike IgG in adult COVID patients and vaccinees shared the same trends in sialylation, galactosylation, and bisection. Namely, spike IgG in adult COVID patients and vaccinees were generally more sialylated and galactosylated and less bisected than bulk IgG from the same groups or healthy controls. Similarly, among pediatric groups, we observed SARS-CoV-2 and MIS-C patients’ spike IgG glycans were on average more sialylated and galactosylated and less bisected than bulk IgG glycans from the same groups or healthy controls, though these differences did not always reach significance (**Fig. 4**).

Between acute COVID and convalescence, the proportion of afucosylated, sialylated, and galactosylated spike IgG significantly decreased in severe COVID patients who received placebo (**Fig. 5**). Similar decreases were also observed for bisected spike IgG and among severe COVID patients who received remdesivir, but these changes did not reach significance. In moderate COVID patients, the same trends were observed but were not significant (**Fig. S7**), possibly due to the smaller sample number in the moderate groups. The one exception was that in moderate COVID patients treated with remdesivir, galactosylated spike IgG abundance trended toward a slight increase between acute disease and convalescence. One possible explanation for this difference is that spike IgG were not as galactosylated in this group during the acute phase as spike IgG in severe COVID patients or moderate COVID patients receiving placebo (**Fig. S7**).

**Figure 5:**
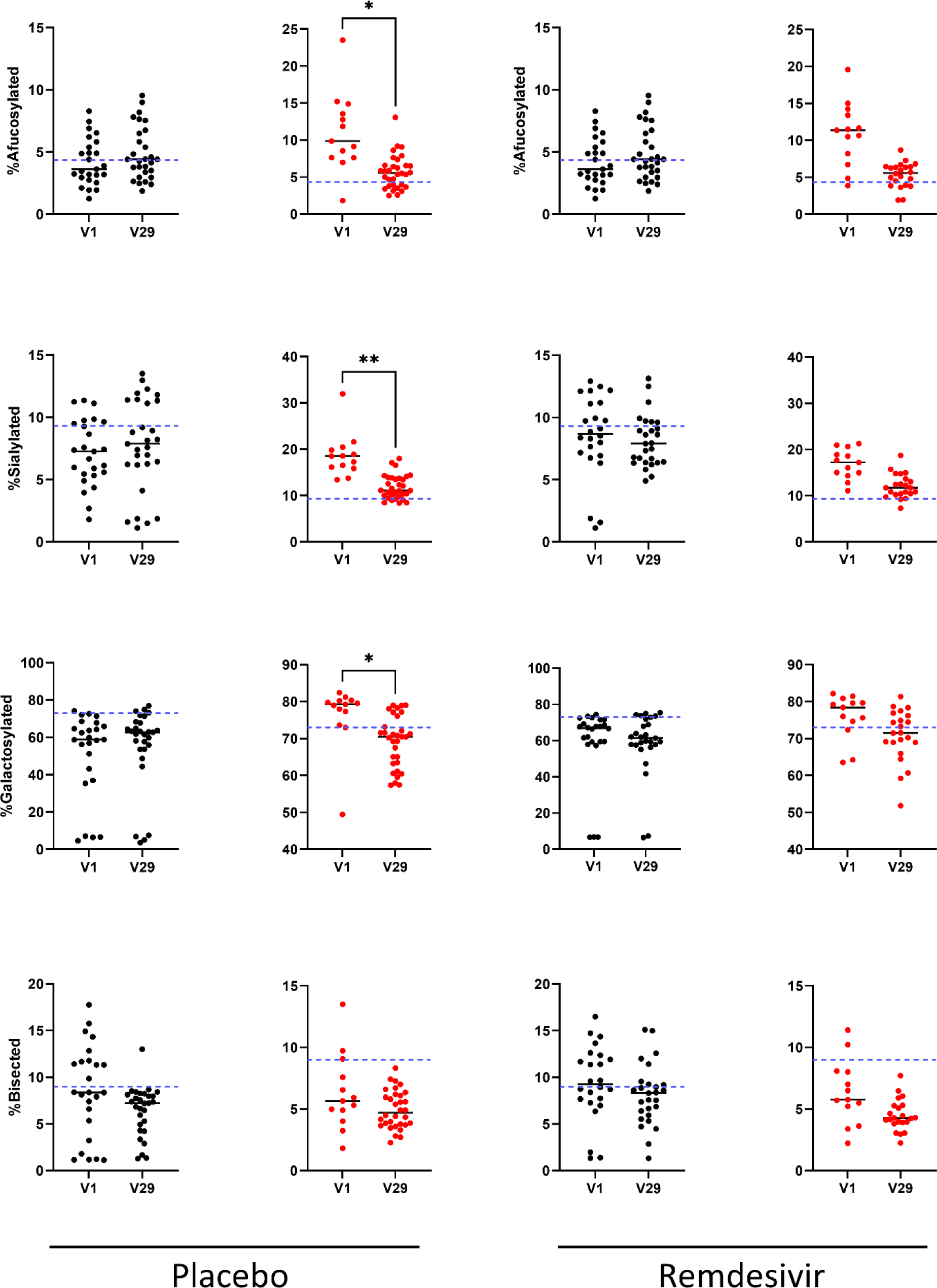
Changes in IgG Fc-glycans between acute infection and convalescence in <u>severe</u> COVID patients. Differences in afucosylated, sialylated, galacosylated, or bisected bulk (black) or anti-spike (red) IgG Fc glycan abundances between acute infection and convalescence in paired serum samples of adults with severe COVID (ordinal scale [OS] 5 and 7), who received either remdesivir or placebo. Wilcoxon matched-pairs signed rank test (* p<0.05, ** p<0.01, *** p<0.001, **** p < 0.0001)

In general, there was less change in bulk IgG Fc glycosylation between acute COVID and convalescence in all adult COVID groups (**Fig. 5, Fig. S7**). There were no differences in overall glycan abundance levels between adult male and female groups during acute SARS-CoV-2 infection, apart from higher bulk IgG Fc galactosylation in male patients compared to female patients (**Fig. S8**).

There were also no significant correlations between oropharyngeal viral load as determined by RT-PCR[40] and bulk or spike IgG Fc glycosylation (**Fig. S9**). Sialylation (in bulk IgG) and galactosylation levels (in both bulk and spike IgG) significantly decreased with age, whereas both bulk and spike IgG Fc bisection levels increased with age (**Fig. S9**).

We observed different trends in spike IgG Fc glycosylation between baseline and seven days post-second dose in vaccinees, namely, no change in sialylation, significantly increased galactosylation, and significantly decreased bisection following vaccination (**Fig. S10**). At the same time, we found a similar significant decrease in spike IgG Fc afucosylation between baseline and seven days post-second dose in vaccinees as observed between acute and convalescent COVID, though the baseline abundance of afucosylated spike IgG in vaccinees was lower than that in acute COVID.

### Correlation Analysis

We then evaluated relationships between spike IgG glycan abundances and inflammatory cytokine/chemokine levels (**Fig. 6)** We observed a positive correlation between bisected spike IgG abundances and IL-6 levels during acute moderate COVID in adults. During acute severe COVID in adults, we observed a negative correlation between galactosylated spike IgG abundances and IL-6 levels and a positive correlation between bisected spike IgG abundances and IFNγ levels. In vaccinees, both afucosylated and bisected spike IgG abundances correlated positively with MIP-1α levels. For MIS-C patients, both afucosylated and sialylated spike IgG abundances correlated inversely with TNFα levels. Sialylated spike IgG also correlated inversely with IL-6 levels in MIS-C patients. Finally, bisected spike IgG abundances in MIS-C patients positively correlated with MCP-3 levels.

**Figure 6:**
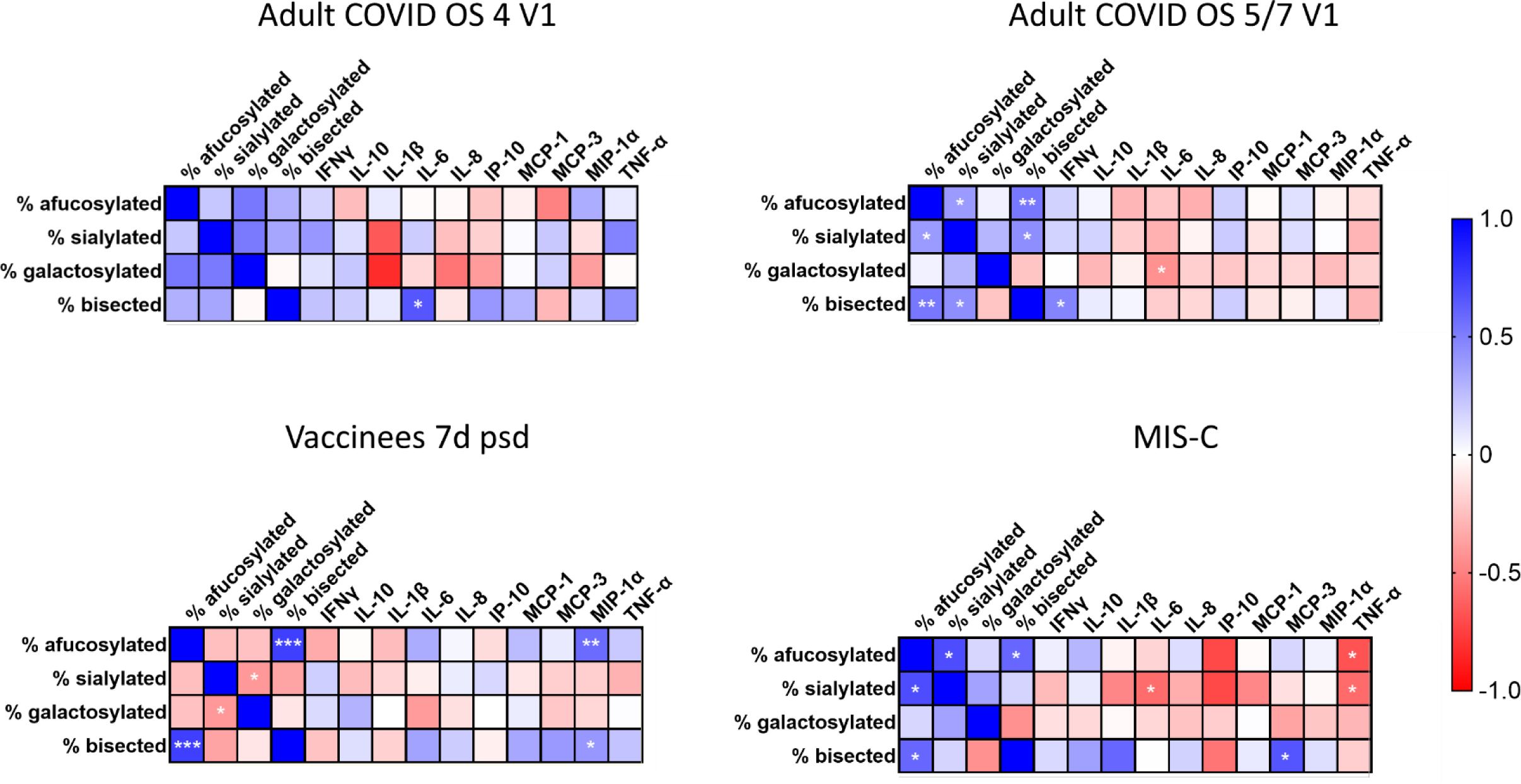
Inverse correlations observed between afucosylated, sialylated, or galactosylated glycan abundances and pro-inflammatory cytokine levels in adult COVID and MIS-C patients are not observed in vaccinees. Spearman correlation coefficients were estimated between serum cytokine/chemokine levels and spike IgG glycan abundances in moderate (OS 4) and severe (OS5/7) COVID at enrollment (V1), mRNA vaccine recipients at 7 days after second vaccine dose (7d psd), and MIS-C patients. Gradient from blue to red indicates strength of positive or inverse correlation, respectively. Significant relationships are indicated by asterisks (* p<0.05, ** p<0.01, *** p<0.001).

## Discussion

This study contributes to a growing body of work aiming to understand how IgG Fc glycosylation impacts antibody effector function, particularly as it relates to COVID immunopathology. All adult COVID patient groups had increased levels of inflammatory cytokines and chemokines in sera during acute COVID compared to healthy controls, as other studies have found[8, 41]. Also similar to other published data[17–19, 21, 23], we found increased proportions of afucosylated spike IgG Fc glycans in COVID patients during acute infection, but not following SARS-CoV-2 mRNA vaccination. While we found significant differences in the abundances of spike IgG Fc glycans with other modifications (i.e., sialylation, galactosylation, bisection), only changes in spike IgG Fc afucosylation seemed to be unique to SARS-CoV-2 infection. For this reason, and as a result of prior *in vitro* and *in vivo* data linking IgG afucosylation with increased affinity for FcγRIIIa and FcγRIIIb[26, 28, 29], increased proinflammatory cytokine secretion[17, 21], increased monocyte and neutrophil recruitment[17], and increased expression of FcγRIIIa on monocytes of patients that progressed to severe COVID[17], spike IgG afucosylation is of particular interest for understanding COVID immunopathology. At the same time, we found both inflammatory cytokine/chemokine levels and spike glycan abundances generally returned to healthy control values at convalescence, signaling a short half-life for these infection-associated changes. Notably, while we and others find the abundance of afucosylated spike IgG declines from acute infection into convalescence and from baseline to post-second vaccine dose[18, 19, 23], not all groups observe this trend[17, 21], which may reflect differences in cohorts or timing of specimen collection. While inflammatory markers broadly correlated with altered spike IgG Fc glycan abundances, our correlation analyses identified only a few significant relationships between cytokine levels and spike glycan abundances in adult acute COVID groups: a negative correlation between galactosylation and IL-6 (OS5/7), and positive correlations between bisection and IL-6 (OS4) and IFNγ (OS5/7). Siekman *et al.* also found significant negative correlations between spike IgG1 Fc galactosylation and IL-1β, IL-6, and IL-8 levels and spike IgG1 Fc sialylation and IL-6 levels[19]. These observations align with the theory that galactosylation is generally anti-inflammatory and bisection is generally pro-inflammatory[25, 32] and suggest that glycosylation modifications may be differentially impacting inflammatory cytokine expression and thus inflammation in COVID. Following vaccination, we found afucosylated and bisected spike IgG abundances correlated with MIP-1α levels, all of which were very low.

Pediatric MIS-C patients also had elevated inflammatory cytokine and chemokine levels in our study. Like adult COVID patients, MIS-C patients had increased proportions of afucosylated spike IgG Fc glycans, which has not been previously published. Similar to adult COVID results, we identified significant correlations between glycosylation patterns and inflammatory markers in paired MIS-C samples, including a negative correlation between afucosylated spike IgG Fc glycans and TNFα, negative correlations between sialylated glycans and TNFα and IL-6, and a positive correlation between bisected glycans and MCP-3. While this is consistent with observed anti-inflammatory activity of IgG Fc sialylation and the proposed inflammatory activity of bisection[25, 33, 34, 42], the negative correlation between spike IgG afucosylation and TNFα does not align with the theory that afucosylation is generally inflammatory. Still, Siekman *et al.* also found a negative relationship between afucosylation and TNFα in adult COVID patients, though in this case it was not statistically significant[19]. Whether these relationships are biologically relevant will require follow up studies. Longitudinal samples prior to the onset of MIS-C as well as after resolution would enable us to determine whether the observed glycosylation patterns precede onset of the condition or return to healthy control levels upon recovery, as has been established with severe adult COVID[17, 19, 23]. Previous work investigating altered IgG glycosylation in MIS-C patients is limited, with our literature searches only finding a paper from Bartsch *et al.*, who found both decreased galactosylation and fucosylation of bulk IgG Fc in severe MIS-C samples compared to pediatric healthy controls[22]. Notably, Bartsch *et al.* did not examine antigen-specific IgG Fc glycosylation in the context of MIS-C as we did here[22]. As part of the same work, Bartsch *et al.* note that severe MIS-C patients had increased monocyte activation[22]. Increased stimulation of FcγRIII by afucosylated antigen-specific IgG may thus be a contributing factor in the systemic inflammatory state characteristic of MIS-C, thus our observations warrant more detailed investigation.

Much of the published research on IgG glycosylation in COVID utilizes mass spectrometry for identifying glycan structure patterns in samples[18–20, 22, 24]. However, this method has some limitations, including sensitivity, throughput, and cost. Overall, analyzing antigen-specific IgG Fc glycosylation by CE worked well for our study, as the method is high-throughput, more cost effective, and highly consistent between separate runs of the same sample in our hands. While results between biological replicates analyzed by both CE and mass spectrometry were also generally consistent, the proportions of galactosylated Fc glycans were found to be higher by CE than by LC-MS/MS. Likely contributing to this discordance was the fact that not all glycans in our panel could be adequately separated by CE[43, 44], and positive glycan identification was limited to the contents of our panel of 17 glycans, though more numerous than the total number of glycoforms reported by other groups using mass spectrometry (n = 14)[18]. We were also unable to generate usable glycan data for 159/282 (56%) spike IgG and 232/341 (68%) bulk IgG samples.

Because of these limitations, it is possible there were differences in spike IgG Fc glycosylation between groups or correlations between glycans and cytokines that were not captured by our data, though our results generally agree with those from similar studies[17–19, 21, 23]. Such inherent limitations of CE for analyzing antibody glycan structures emphasize its utility for observing broad trends in IgG glycosylation patterns using large numbers of samples. Indeed, CE may not be as well-suited for detailed IgG subclass-specific Fc glycosylation analysis as LC-MS/MS, which suffers from lower sensitivity than CE[44], or glycan engineering, which allows for precise control over N297 glycosylation and is our choice for subsequent mechanistic testing[45]. It should be noted that groups utilizing mass spectrometry to analyze Fc glycosylation changes following SARS-CoV-2 infection and mRNA vaccination found similar trends across different IgG subclasses, with the exception of afucosylation of IgG3 after vaccination[17, 18]. An additional limitation of our study is that we only looked at vaccine recipient samples after the second dose of the vaccine, so we may have missed glycosylation changes or cytokine responses present after the priming dose, though other groups observed similar Fc glycosylation trends between the first and second doses[18].

Our findings on the alteration of antigen-specific IgG glycosylation patterns following SARS-CoV-2 infection or vaccination corroborate existing studies. Regarding MIS-C, however, our results point towards previously underexplored relationships that may be impacting the disease. Our observations warrant further investigation, which could include considerations of IgG titer, subclass, and temporal analysis of spike IgG Fc glycosylation over MIS-C and COVID disease course, including in long-COVID. Additional *in vitro* and *in vivo* work using glycan-engineered monoclonal antibodies with specific glycan modifications to stimulate monocytes or neutrophils for assessment of inflammatory cytokine/chemokine secretion might further elucidate the contributions of afucosylation, galactosylation, bisection, and sialylation to these effector functions. As patterns of antigen-specific IgG Fc afucosylation seem to be consistent across diverse viral infections, such studies could have broad applicability for understanding drivers of inflammation during viral infections[23, 31].

## Data Availability

All data produced in the present study are available upon reasonable request to the authors.

## Author Contributions

Jacob D. Sherman1,^2,3,4,5^, Vinit Karmali^3^, Bhoj Kumar^3,5^, Trevor W. Simon^2,5^, Sarah Bechnak^3,4^, Anusha Panjwani^3,4^, Caroline R. Ciric^3,4^, Dongli Wang^4^, Chris Huerta^4^, Brandi Johnson^4,6^, Evan J. Anderson^5^, Nadine Rouphael^5^, Matthew H. Collins^5^, Christina A. Rostad^3,5^, Parastoo Azadi^5,7^, Erin M. Scherer^1,2,3,5,6,7,8^

^1^ Writing – original draft

^2^ Method development

^3^ Data collection/analysis

^4^ Sample collection/processing

^5^ Writing – Reviewing and Editing

^6^ Supervision/Project Administration

^7^ Funding acquisition

^8^ Study conception

## Acknowledgments

We thank Ryan McCool and Cory Acreman from the McLellan Lab (University of Texas, Austin) for providing the SARS-CoV-2 HexaPro spike[46]. We also thank Natalia Kozak-Muiznieks, Melisa Willby, and Lesley McGee from the Center for Disease Control and Prevention for generously allowing our use of their ABI3130XL and ABI3500XL and for their support with using the equipment. We thank Teresa Snyder-Leiby from SoftGenetics LLC for her guidance in using the GeneMarker program for our glycan data analysis. We also thank Yongxian Xu for cytokine data analysis, and Daniel Espinoza for sample processing and management. Additionally, we thank Camila Ornelas and Michael Zianni from ThermoFisher Scientific for CE troubleshooting and protocol support.

Supported by the Infectious Diseases Clinical Research Consortium through the National Institute for Allergy and Infectious Diseases of the National Institutes of Health, under award number UM1AI148684 as an IDCRC Early Career Investigator Pilot Award to EMS. The content is solely the responsibility of the authors and does not necessarily represent the official views of the National Institutes of Health.

Mass spectrometry-based glycomics analysis was performed at the Complex Carbohydrate Research Center and was supported in part by the National Institutes of Health (NIH)-funded R24 grant (R24GM137782) to Parastoo Azadi.

This work used samples and data from the Adaptive Covid-19 Treatment Trial (ACTT-1) trial (DOI:10.1056/NEJMoa2007764). The ACTT-1 trial was sponsored and primarily funded by the National Institute of Allergy and Infectious Diseases (NIAID), National Institutes of Health (NIH), Bethesda, MD. This trial has been funded in part with federal funds from the NIAID and the National Cancer Institute, NIH, under contract HHSN261200800001E 75N910D00024, task order number 75N91019F00130/75N91020F00010, and by the Department of Defense, Defense Health Program. This trial has been supported in part by the NIAID of the NIH under award numbers UM1AI148684, UM1AI148576, UM1AI148573, UM1AI148575, UM1AI148452, UM1AI148685, UM1AI148450, and UM1AI148689. The trial has also been funded in part by the governments of Denmark, Japan, Mexico, and Singapore. The trial site in South Korea received funding from the Seoul National University Hospital. Support for the London International Coordinating Centre was also provided by the United Kingdom Medical Research Council (MRC_UU_12023/23). The sites and investigators involved with collection of the samples during the ACTT-1 trial are noted in the original manuscript.

