## Supplemental Materials for "Altered spike IgG Fc N-linked glycans are associated with hyperinflammatory state in adult COVID and Multisystem Inflammatory Syndrome in Children"

### Materials and Methods

#### *Samples*

Secondary research samples were obtained from the NIH-funded ACTT-1 clinical trial[1] from 109 participants with acute COVID from Study Visit 1 (enrollment; occurring ≤12 days post-symptom onset) and Visit 29 (n=218 serum samples). Baseline disease severity by ordinal scale was 4 (moderate; n=29 participants), 5 (severe; n=40 participants), and 7 (critical; n=40 participants). Subjects received either remdesivir treatment (n=57) or a placebo (n=52). A breakdown of the study groups is shown in **Supplementary Table 1**. For all prospective samples, informed consent was obtained prior to study procedures and all studies were Emory University institutional review board (IRB) approved.

For cytokine and IgG glycosylation analysis, unmatched adult healthy control serum samples (n=10) were collected prior to 2019 at Emory University (IRB00103363). For the anti-SARS-CoV-2 spike IgG screening ELISAs, a separate set of unmatched adult healthy control serum samples (n=6) were used (IRB00045821).

Serum and plasma samples were collected at Emory (IRB00045821 and IRB00002061) from 37 healthy adults who received either Pfizer-BioNTech BNT162b2 SARS-CoV-2 vaccine (n=18) or Moderna mRNA-1273 SARS-CoV-2 vaccine (n=19) as part of standard care. Samples were collected at baseline or between six and fifteen days post-second dose ("7 days psd" samples, n=74 samples).

Pediatric serum samples were collected from patients with COVID (n=9) or MIS-C (n=20) at Emory (STUDY00000723), as well as from pediatric healthy controls (n=10) (IRB00087446).

Use of the above secondary research samples for this study was approved by the Emory IRB (STUDY00002583).

*Cytokine/Chemokine Multiplex ELISA*

Serum and plasma samples were heat inactivated in a 56°C water bath for 30 minutes. Cytokine and chemokine levels in 25 µL sera were quantified in duplicate using a custom Mesoscale Discovery (MSD) U-plex assay from the Human Biomarker Group 1 Panel with Human Proinflammatory Panel 1 Controls (MSD C4049-1) on each plate. Cytokine/chemokine data were excluded from analysis if they fell below the quantifiable range.

*Anti-SARS-CoV-2 Spike IgG Screening ELISAs*

Adult COVID and vaccinee baseline samples, as well as all pediatric samples, were screened by SARS-CoV-2 spike IgG ELISA as previously described[2]. Samples were tested in duplicate at a 1:30 and 1:120 dilution. A cutoff was determined for each dilution using six healthy control samples screened at a 1:30 and 1:120 dilution as previously described[3]. Samples were considered seronegative if the average $A_{405}$ at both dilutions fell below the cutoff value. Samples seronegative for anti-SARS-CoV-2 spike IgG were omitted from anti-SARS-CoV-2 spike IgG Fc analysis.

*IgG Purification*

Serum or plasma samples (200 µL[4]) were first desalted using Zeba Spin Desalting plates (ThermoFisher) following manufacturer recommendations. Bulk IgG was then purified from the desalted samples using Melon Gel Spin plates (ThermoFisher) following manufacturer recommendations, excepting the sample incubation step which was extended to 30 minutes at 800 RPM on a plate shaker to improve yield and purity.

*Bulk IgG IdeZ Digest*

Twenty-two µL from each purified bulk IgG sample were separated into $F(ab)_2$ and Fc fragments by IdeZ Protease (New England Biolabs) digestion following manufacturer recommendations with the following alterations: 40 units of IdeZ protease was used per reaction following protocol development; the IdeZ protease and Glycobuffer 2 (10x) (New England Biolabs) were first prepared as a master mix before adding 3 µL to each sample; the incubation at $37^oC$ was extended to 2 hours in a thermocycler. Reactions were prepared in a 96-well PCR plate (ThermoFisher AB-0800-L).

*Bulk IgG Fc Enrichment*

Prior to the addition of bulk IgG digests containing Fc domains, 50 µL of Pierce Protein G Dynabeads (ThermoFisher) were dispensed into each well of a polypropylene U-bottom 96-well plate and collected with a plate magnet (Invitrogen 12027); storage solution was removed, and beads were washed with 100 µL PBS with 0.05% Tween-20 (PBS-T). The wash was removed, and bulk IgG digests were transferred to the washed Protein G Dynabeads and mixed, then the plate was incubated for 30 minutes on a plate shaker at 800 RPM. The supernatant was discarded, and the beads were washed 3x with PBS. IgG Fc was eluted by adding 25 µL 0.1 M citric acid (pH 3.0), followed by incubation for 10 minutes on a plate shaker at 1000 RPM. The eluates were transferred to a 96-well PCR plate containing 4 µL neutralization buffer (1 M sodium carbonate-bicarbonate buffer, pH 10.8).

*Anti-SARS-CoV-2 Spike IgG Isolation and IdeZ Digest*

Fifty µL His-tag Dynabeads (Invitrogen 10104D) were coated with 12 µg His-tagged recombinant SARS-CoV-2 spike in the following manner: 16.5 mL Dynabeads were added to a 50-mL conical tube, and the Invitrogen plate magnet was used to collect the beads. The supernatant was discarded, and 33 mL SARS-CoV-2 spike (0.12 mg/mL in PBS) was added, and the beads were mixed thoroughly before 10 minutes incubation on a rotational mixer. The supernatant was discarded, and the bead-protein complex was washed 4x with PBS, with thorough mixing between each wash. The bead-protein complex was resuspended in 33 mL PBS-T, which was divided into 100 µL per well of V-bottom 96-well plates.

Anti-SARS-CoV-2 spike IgG enrichment was done in three rounds, with each round proceeding in the following manner: Remaining bulk IgG samples were applied to the Dynabead-spike complexes and mixed thoroughly before 30 minutes incubation on a plate shaker at 800 RPM. The beads were collected with a plate magnet and the supernatants were either transferred to the next set of beads (for the 2$^{nd}$ and 3$^{rd}$ enrichments) or discarded (after the 3$^{rd}$ enrichment). The beads were then washed 3x with PBS-T with 40 mM imidazole, followed by one wash with PBS. After enrichment, beads from each round of enrichment of the same sample were pooled into a 96-well PCR plate for IdeZ digest.

IdeZ digestion was performed according to manufacturer recommendations with 80 units of IdeZ per reaction and the following alterations: the Glycobuffer 2 (1x) volume was doubled for a 50 µL total reaction volume; the reagents (Glycobuffer 2 [10x], molecular biology grade water, and IdeZ) were assembled as a master mix prior to addition to the samples, and the digestion was performed directly on the Dynabead-spike-IgG complexes for 2 hours at 37$^{o}$C on a plate shaker set to 800 RPM. After digestion, the beads were collected with a plate magnet and the 50 µL supernatants transferred to a 96-well PCR plate for rapid PNGaseF digest.

*Rapid PNGaseF Digest*

Asparagine(N)-linked deglycosylation of bulk or spike IgG Fc were performed using Rapid PNGaseF (New England Biolabs) according to manufacturer recommendations with the following alterations: a sufficient volume of Rapid PNGase F Buffer (5x) was used to bring the final buffer concentration to 1x, 1 µL enzyme was used per reaction, and incubation was performed for 30 minutes at 50$^{o}$C in an Eppendorf Mastercycler EP Gradient Thermocycler (model 5341). Reactions were performed in a 96-well PCR plate (ThermoFisher AB-0800-L). After incubation, the completed reactions were transferred to a polypropylene 96-well V-bottom plate and centrifuged at 2000 xg for 5 minutes, then allowed to dry completely overnight inside of a biosafety cabinet. Once dried, the plate was again centrifuged at 2000 xg for 5 minutes, then sealed and stored at 4$^{o}$C until use for fluorescent labelling.

*Fluorescent Labelling of IgG Fc Glycans*

Dried PNGaseF deglycosylation products were reconstituted in 4 µL molecular biology grade water. A solution of 20 mM 8-aminopyrene trisodium salt (APTS) (Sigma-Aldrich 09341) and 3.6 M citric acid (Sigma-Aldrich 27487) was prepared and mixed in equal volume with 200 mM 2-picoline borane (Sigma-Aldrich 654213) in dimethyl sulfoxide (Sigma-Aldrich 08418). From this point forward, samples and reagents were protected from light as much as possible. The reaction was conducted by adding 10 µL of the labelling solution to each reconstituted sample, mixing thoroughly, and transferring to a 96-well PCR plate (ThermoFisher AB-0800-L). The reactions were incubated in a thermocycler at 37$^{o}$C for 16 hours, then quenched with 125 µL 20% reverse-osmosis purified water, 80% acetonitrile (Sigma-Aldrich 34851) (v/v).

*HILIC-SPE Purification of APTS-N-Glycans*

HILIC-SPE was performed to isolate APTS-labelled N-glycans from other reagents as described previously [5], with the following modifications: an 0.45 um wwPTFE membrane plate (Pall 8684) was used; APTS-sample/BioGel P10 incubation was extended to 10 minutes; the wash volumes (the 20% water, 80% acetonitrile (v/v), 100 mM triethylamine (Sigma-Aldrich 81101) wash; and the 20% water, 80% acetonitrile (v/v) wash) were increased to 600 µL; after initial incubation with 100 µL molecular biology grade water, elution was performed by the single addition of 100 µL water, followed by collection by centrifugation at 1000xg for 2 minutes into a V-bottom polypropylene plate. The entire protocol was performed with care to minimize light exposure. APTS-labeled N-glycan samples were stored at -20$^{o}$C until analysis by capillary electrophoresis (CE).

*Capillary Electrophoresis*

Agilent APTS-labeled glycan reference standards were initially reconstituted in 100 µL molecular biology grade water, protected from light, and stored at -20$^{o}$C until use. Bracketing standard (GKSP-500), G0 (GKSP-301), G1 (GKSP-317), G2 (GKSP-304), G0F-N (GKSP-402), G0-N (GKSP-401), and G0F (GKSP-302) were diluted 1/80 before use, and the IgG N-Glycan Library (GKSP-005) was diluted 1/15 before use. Dilutions were made in molecular biology grade water immediately prior to sample preparation.

After all components were thawed at room temperature, a glycan analysis master mix was prepared corresponding to 0.125 µl GeneScan 500 LIZ size standard (Applied Biosystems 4322682), 9.875 µL Hi-Di formamide (Applied Biosystems 4401457), and 3 µL bracketing standard per sample. The master mix was dispensed as 13 µL into each well of a 96-well polypropylene PCR plate (Applied Biosystems N8010560), and 10 µL of each APTS-labeled N-glycan sample was added. The N-glycan reference panel was prepared by combining 3 µL each of, G0, G1, G2, G0F-N, G0-N, and G0F with 13 µL sequencing master mix. The IgG N-glycan reference standard was added as a 3 µL sample to a separate well with 13 µL sequencing master mix. Sample volume was adjusted, if necessary, according to signal strength on subsequent runs.

APTS-labeled N-Glycan samples were analyzed using an ABI3130XL or ABI3500XL instrument with 50 cm capillaries containing POP-7 polymer (Applied Biosystems). When run on the ABI3130XL, samples were injected at 1.6 kV for 24 seconds, then fluorescence data was collected for 1800 seconds at 15 kV, and the oven temperature was set to 60$^{o}$C. When run on the ABI3500XL, the run voltage was 19.5 kV, data was collected for 1330 seconds, and all other parameters remained the same.

APTS-labeled N-linked glycan electropherograms were analyzed using GeneMarker (v3.0.1). Traces were aligned to the internal bracketing standard peaks and G0F peak using the macromolecule tool. A panel was constructed using the pooled glycan standard and IgG N-glycan library traces and aligned to APTS-labeled samples to identify glycan peaks. Area-under-the-curve (AUC) was quantified for each peak in the linear range of the ABI3130XL (100-7000 RFU) to determine glycoform abundance out of total glycan AUC. Glycans were grouped based on the presence or absence of modifying residues (galactosylation, sialylation, bisection, and afucosylation). As four glycans in our panel did not adequately separate by CE, and some peaks were present in samples that were not present in our standard panels, analyses were done conservatively by taking the mean of the maximum (assuming all unknowns fit into the category) and minimum (assuming all unknowns did not fit into the category).

*Mass Spectrometry*

The IgG Fc sample (5 µg at 0.234 mg/mL) was desalted with a 3 kDa MWCO filter and further reduced and alkylated with 5 mM DTT and 10 mM iodoacetamide. The proteins were then digested with a 1:20 ratio of trypsin at 37oC for 18 hours. The resulting peptides were subjected to LC-MS/MS on an Orbitrap Eclipse mass spectrometer (ThermoFisher) equipped with 3000 RSLCnano system. In positive ion mode, a 120 000 resolution full mass scan was collected, followed by data-dependent MS/MS (higher collision energy (HCD) fragmentation; 30,000 resolution) scans of the highest abundance peaks within a 3 sec time frame. Data were processed with Byonic software (Protein Metrics; v4.0.12) and searched against the IgG Fc sequence and a catalogue of more than 59 human N-linked glycans. The precursor mass tolerance and fragment mass tolerances were set to 5 ppm and 10 ppm, respectively. Additional modifications including deamidation of asparagine and glutamine, carboxymethylation of cysteine, and oxidation of methionine were included in the search. Byonic software was used to assist in the characterization of these glycosylation sites and to confirm the non-glycosylated ones. These assignments were based mainly on delta mass of the assumed glycopeptide, characteristic oxonium ions and neutral losses following HCD fragmentation, and y and b ions along the peptide backbone as previously described[6]. The relative percentages of each glycoform were determined by deconvolution of the LC-MS data at the full MS level, then determining the AUC for each full MH+

*Statistical Analysis*

All statistical analyses were performed in GraphPad Prism (v9.4.1). Multiple group comparisons for cytokine/chemokine concentrations and glycan abundances were analyzed by Kruskal-Wallis with Dunn's test, and paired data between timepoints were analyzed by Wilcoxon matched-pairs signed rank test. Correlation matrices were generated by Spearman correlation.

**Supplementary Table 1:** *Study groups and samples collected*

| Group | Sub-Group | Number of Samples | Timepoints |
|---|---|---|---|
| Adult COVID samples from ACTT-1 Study[1] | Moderate (ordinal scale 4), Placebo | 12 | Visits 1 (enrollment) and 29 |
| | Moderate (ordinal scale 4), Remdesivir Treated | 17 | Visits 1 (enrollment) and 29 |
| | Severe (ordinal scale 5), Placebo | 20 | Visits 1 (enrollment) and 29 |
| | Severe (ordinal scale 5), Remdesivir Treated | 20 | Visits 1 (enrollment) and 29 |
| | Critical (ordinal scale 7), Placebo | 20 | Visits 1 (enrollment) and 29 |
| | Critical (ordinal scale 7), Remdesivir Treated | 20 | Visits 1 (enrollment) and 29 |
| Adult SARS-CoV-2 mRNA vaccinee samples | mRNA-1273 | 19 | Baseline and 7 days post-second dose |
| | BNT162b2 | 18 | Baseline and 7 days post-second dose |
| Pediatric COVID and MIS-C samples | Acute COVID | 9 | Enrollment |
| | MIS-C | 20 | Enrollment |
| Healthy Controls | Pediatric | 10 | Enrollment |
| | Adult | 10 | Enrollment |
| | Adult | 6 (for anti-S-trimer IgG screens) | Enrollment |

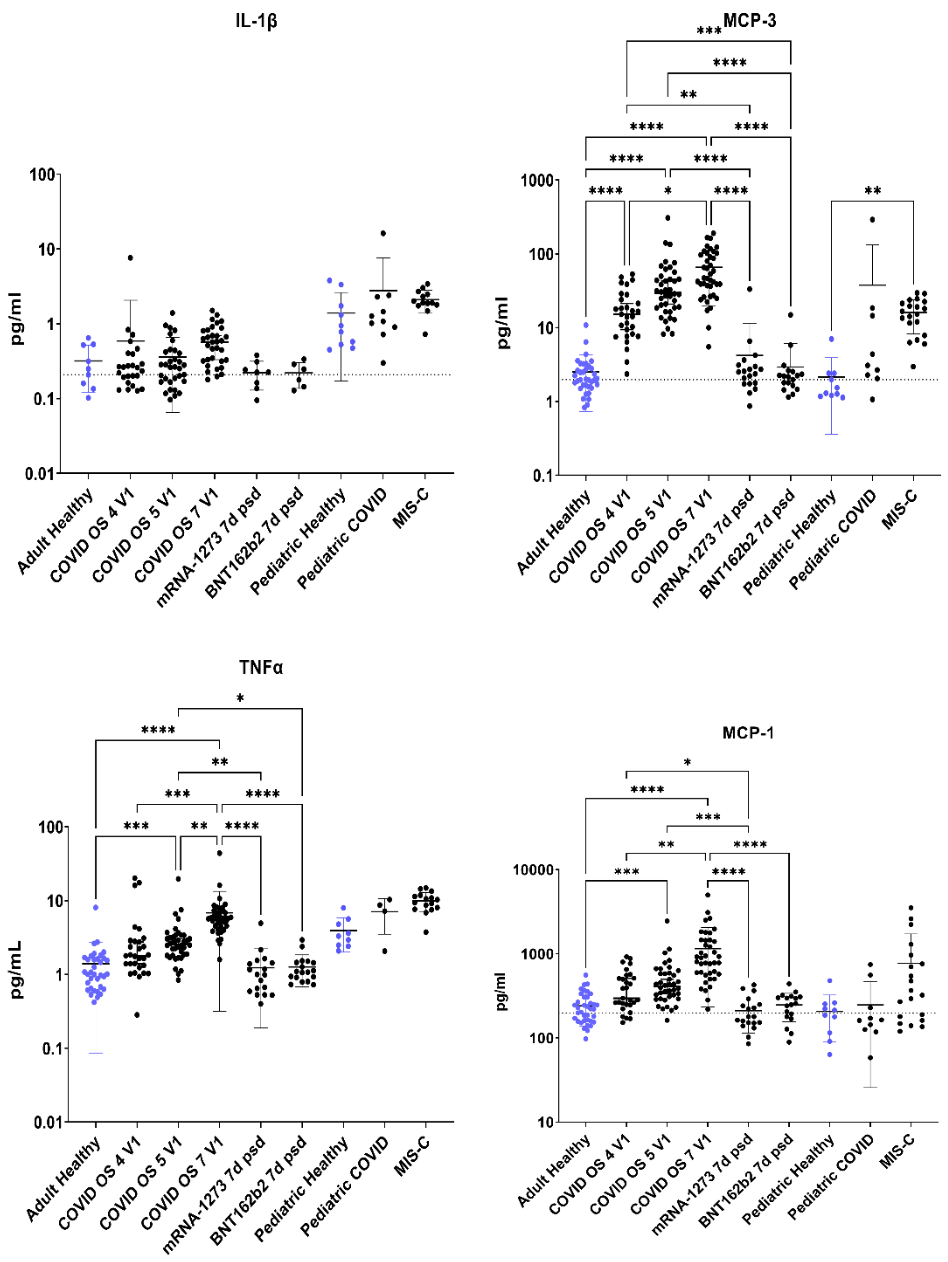


**Figure S1: Proinflammatory cytokines and chemokines are significantly elevated in adult COVID patients compared to healthy controls (related to Fig. 1).** Differences in IL-1β, MCP-1, MCP-3, and TNF levels between adult healthy controls, which includes vaccinees at baseline, patients enrolled in ACTT-1 with different COVID severities by ordinal scale (OS) at enrollment (V1), or vaccinees (7 days post-second dose, 7d psd); as well as between pediatric healthy controls or pediatric patients with SARS-CoV-2 or MIS-C. Kruskal-Wallis with Dunn's test (* $p<0.05$, ** $p<0.01$, *** $p<0.001$, **** $p < 0.0001$)

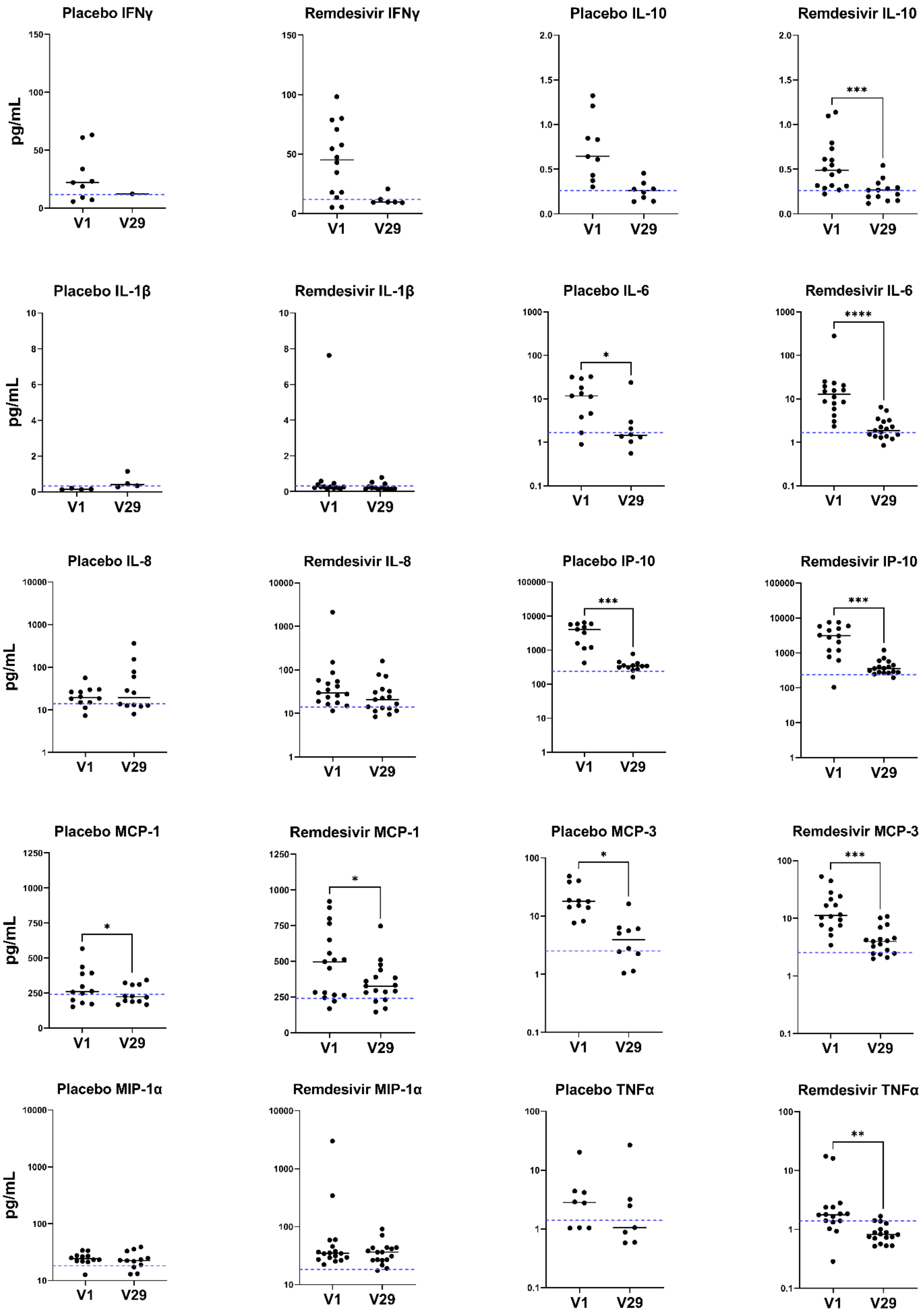


**Figure S2: Proinflammatory cytokines and chemokines are significantly decreased in adult moderate COVID between acute infection and convalescence.** Differences in cytokine and chemokine levels between acute infection and convalescence in paired serum samples of adults with moderate COVID (ordinal scale [OS] 4), who received either remdesivir or placebo. Healthy control mean value indicated by dashed blue line. Wilcoxon matched-pairs signed rank test (* $p<0.05$, ** $p<0.01$, *** $p<0.001$, **** $p < 0.0001$) Plots without significance bars lacked sufficient paired data for test.

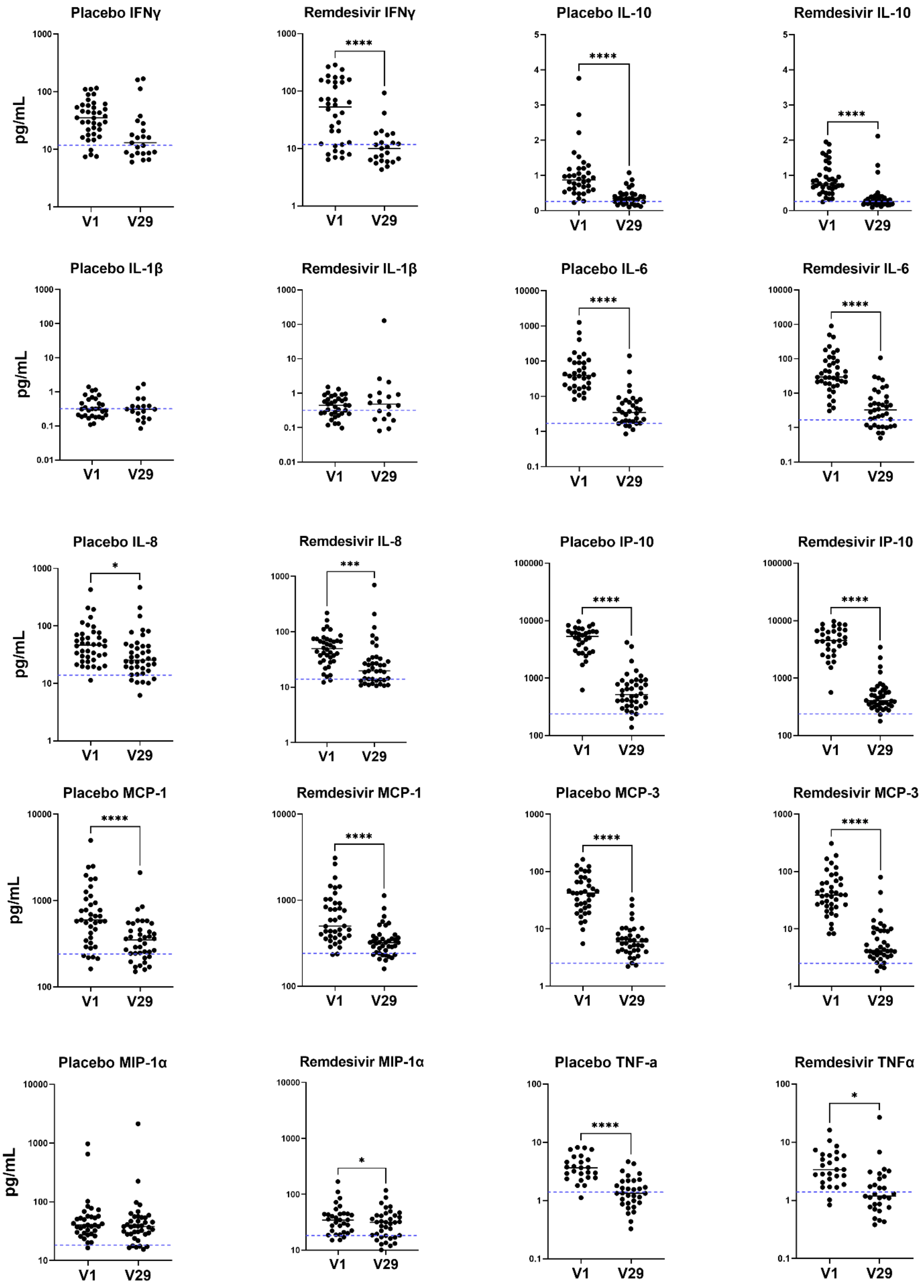


**Figure S3: Proinflammatory cytokines and chemokines are significantly decreased in adult severe COVID between acute infection and convalescence.** Differences in cytokine and chemokine levels between acute infection and convalescence in paired serum samples of adults with severe COVID (ordinal scale [OS] 5 and 7), who received either remdesivir or placebo. Healthy control mean value indicated by dashed blue line. Wilcoxon matched-pairs signed rank test (* $p<0.05$, ** $p<0.01$, *** $p<0.001$, **** $p < 0.0001$) Plots without significance bars lacked sufficient paired data for test.

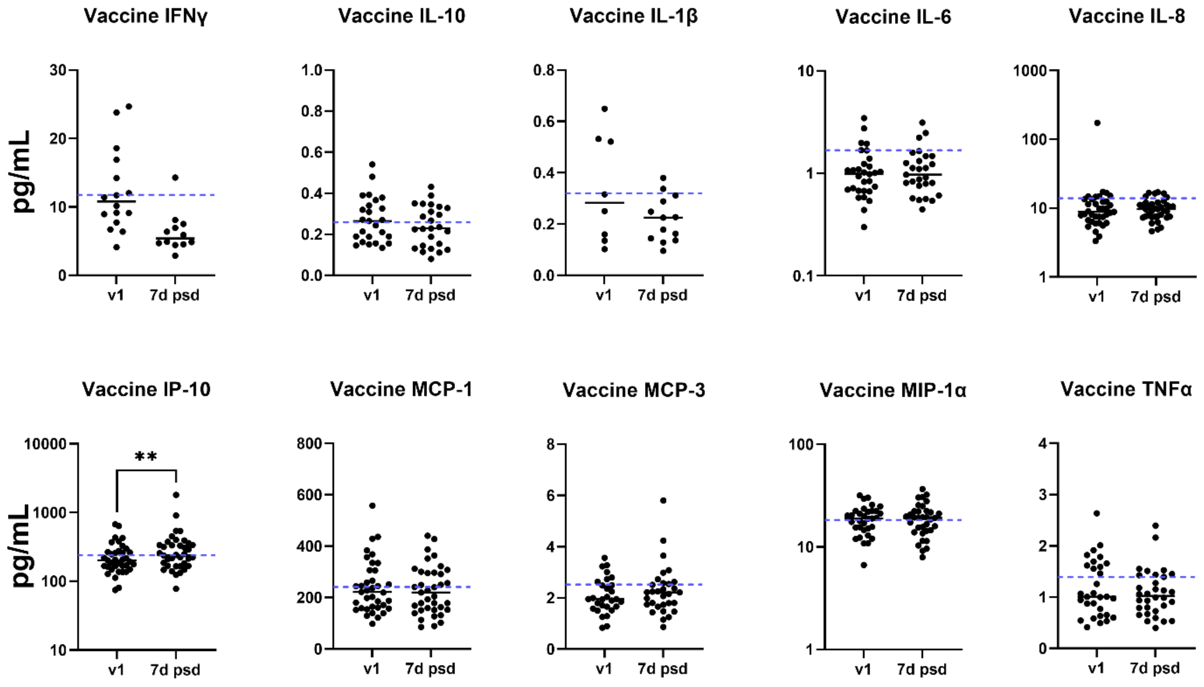


**Figure S4: Proinflammatory cytokines and chemokines are mostly unchanged pre- and post-vaccination in adult mRNA COVID vaccine recipients.** Differences between cytokine and chemokine levels pre-vaccination and 7 days post second dose (7d psd) in paired serum samples of adults who received a COVID mRNA-based vaccine. Healthy control mean value indicated by dashed blue line. Wilcoxon matched-pairs signed rank test (* $p<0.05$, ** $p<0.01$, *** $p<0.001$, **** $p < 0.0001$) Plots without significance bars lacked enough sufficient data for test.

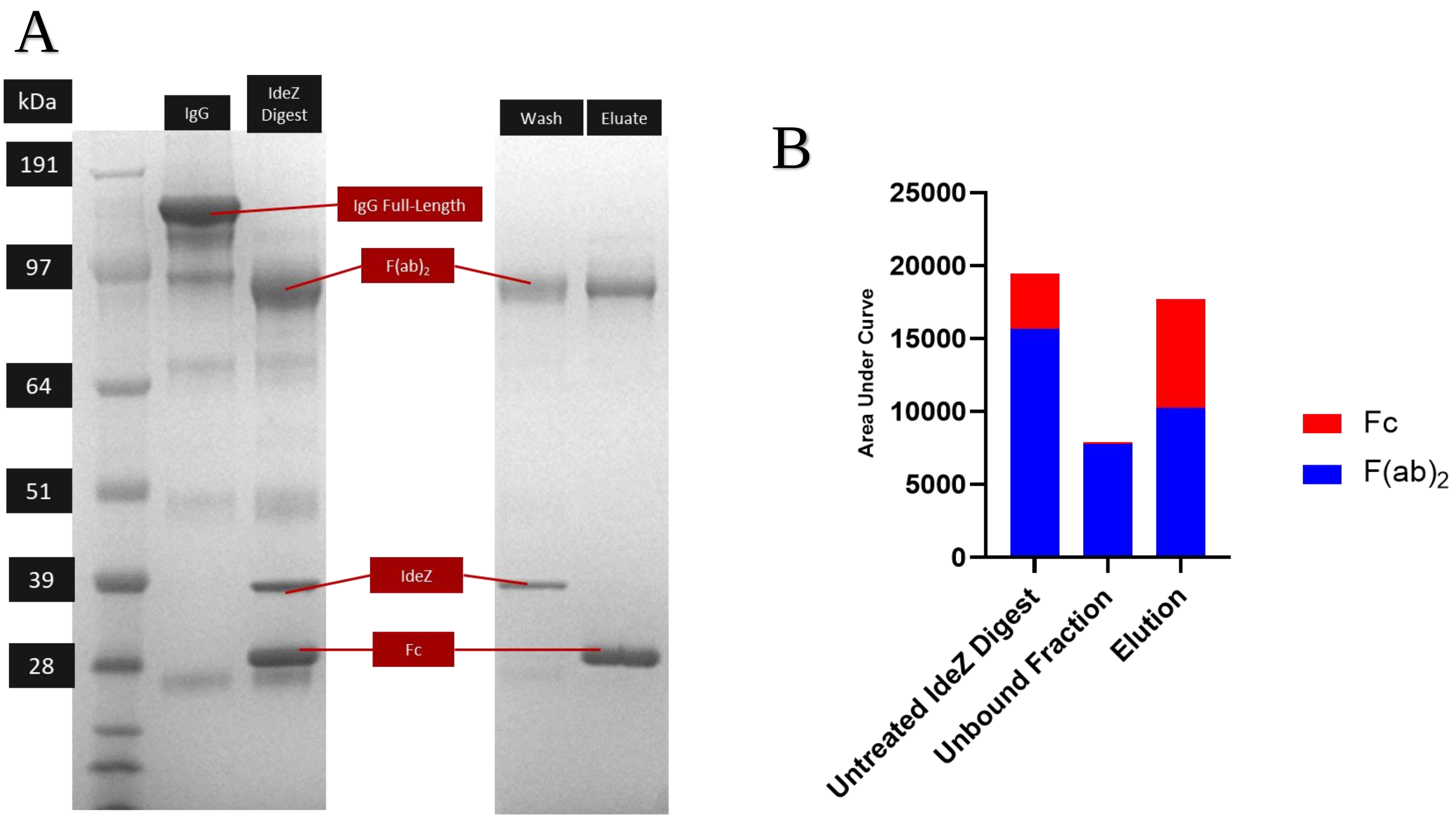


**Figure S5: Protein G beads enrich both F(ab)$_2$ and Fc domains from bulk IgG Fc IdeZ digests. A)** 20 µg bulk IgG purified from serum was digested with 40 units IdeZ in solution for 2 hours, then treated with protein G beads, washed, and eluted with 0.1 M citric acid pH 3.0. Samples of bulk IgG, untreated IdeZ digest, wash fraction, and eluate were analyzed by SDS-PAGE with reference to a protein molecular weight ladder, and **B)** area-under-the-curve quantified for Fc and F(ab)$_2$ bands by densitometry to estimate the products enriched by protein G beads. The SDS-PAGE image was cropped to omit irrelevant lanes (i.e., alternative wash conditions).

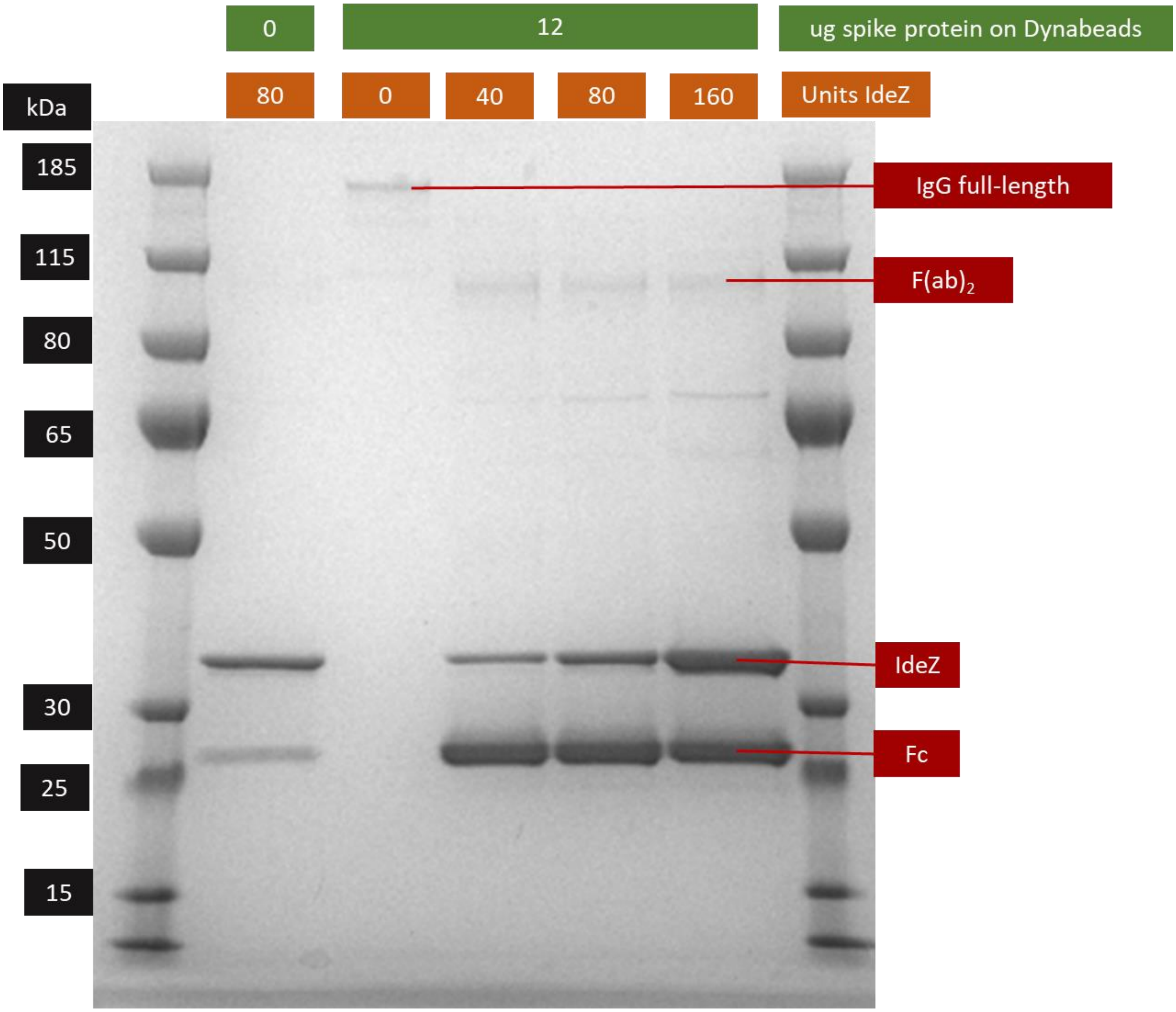


**Figure S6: SDS-PAGE analysis demonstrating IdeZ cleavage of anti-spike IgG Fc from spike-coated Dynabeads.** Bulk IgG purified from 200 µL anti-spike seropositive serum was enriched three times on Dynabeads coated with 0 or 12 µg spike, then digested for 2 hours with 0-160 units IdeZ protease, and the supernatants were analyzed by SDS-PAGE with reference to a protein molecular weight ladder.

**Table S2: Reproducibility of bulk IgG Fc-Glycan analysis by capillary electrophoresis (technical replicates, separate analyses)**

| Sample | %afucosylated (range) | %sialylated (range) | %galactosylated (range) | %bisected (range) |
|---|---|---|---|---|
| 7-Mar-22 | 0.7%-6.5% | 5.8%-22.6% | 56.5%-73.3% | 2.1%-7.9% |
| 15-Mar-22 | 0%-0% | 3.6%-12.9% | 56.4%-65.7% | 0%-0% |
| 22-Apr-22 | 0%-0% | 4.1%-13.9% | 54%-63.7% | 0%-0% |
| **SD of Means** | **2%** | **3%** | **3%** | **3%** |

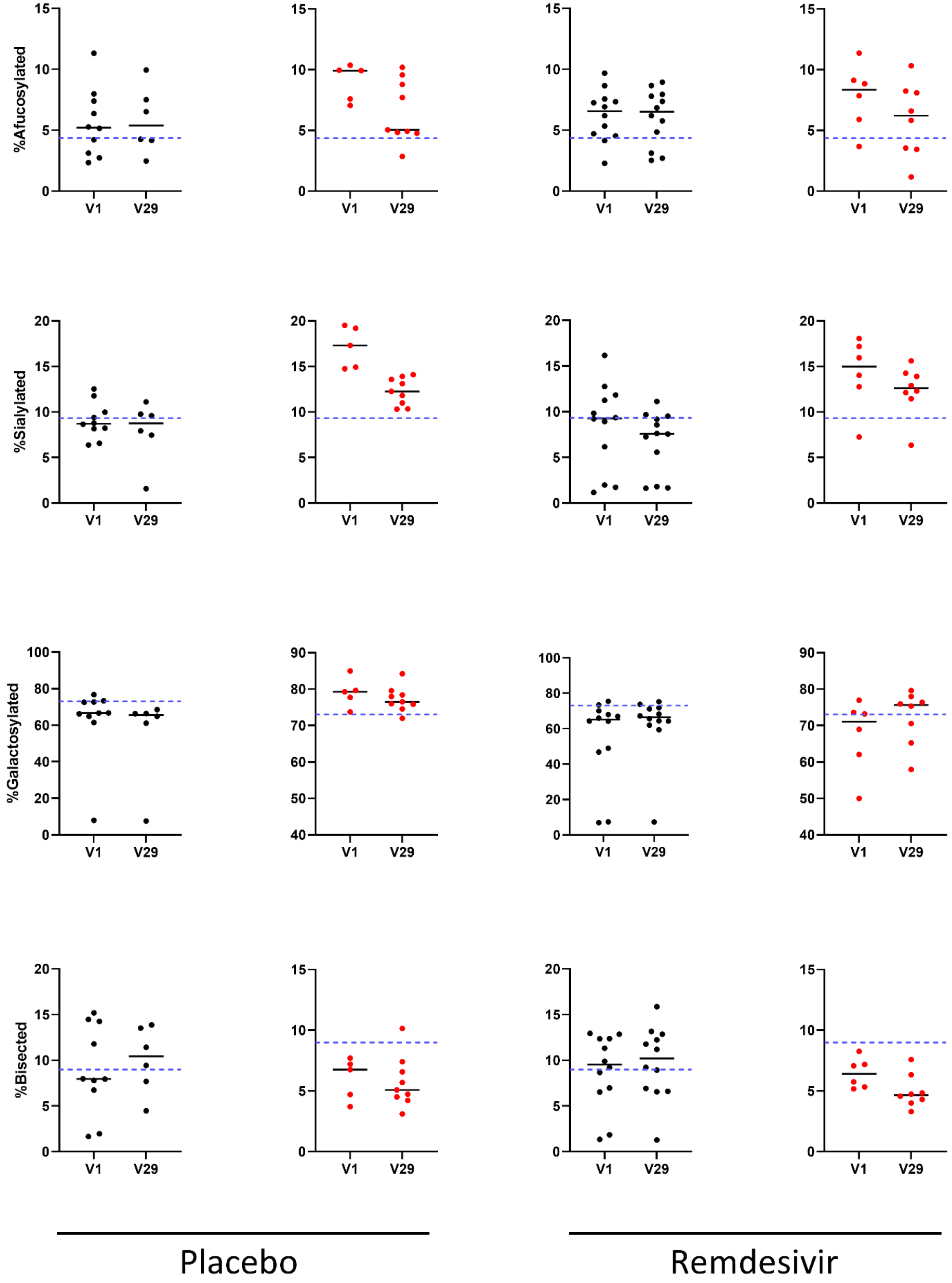


**Figure S7: Changes in IgG Fc-glycans Between acute infection and convalescence in <u>moderate</u> COVID patients.** Differences afucosylated, sialylated, galacosylated, or bisected bulk (black) or anti-spike (red) IgG Fc glycan abundances between acute infection and convalescence in paired serum samples of adults with moderate COVID (ordinal scale [OS] 4), who received either remdesivir or placebo. Wilcoxon matched-pairs signed rank test (* p<0.05, ** p<0.01, *** p<0.001, **** p < 0.0001)

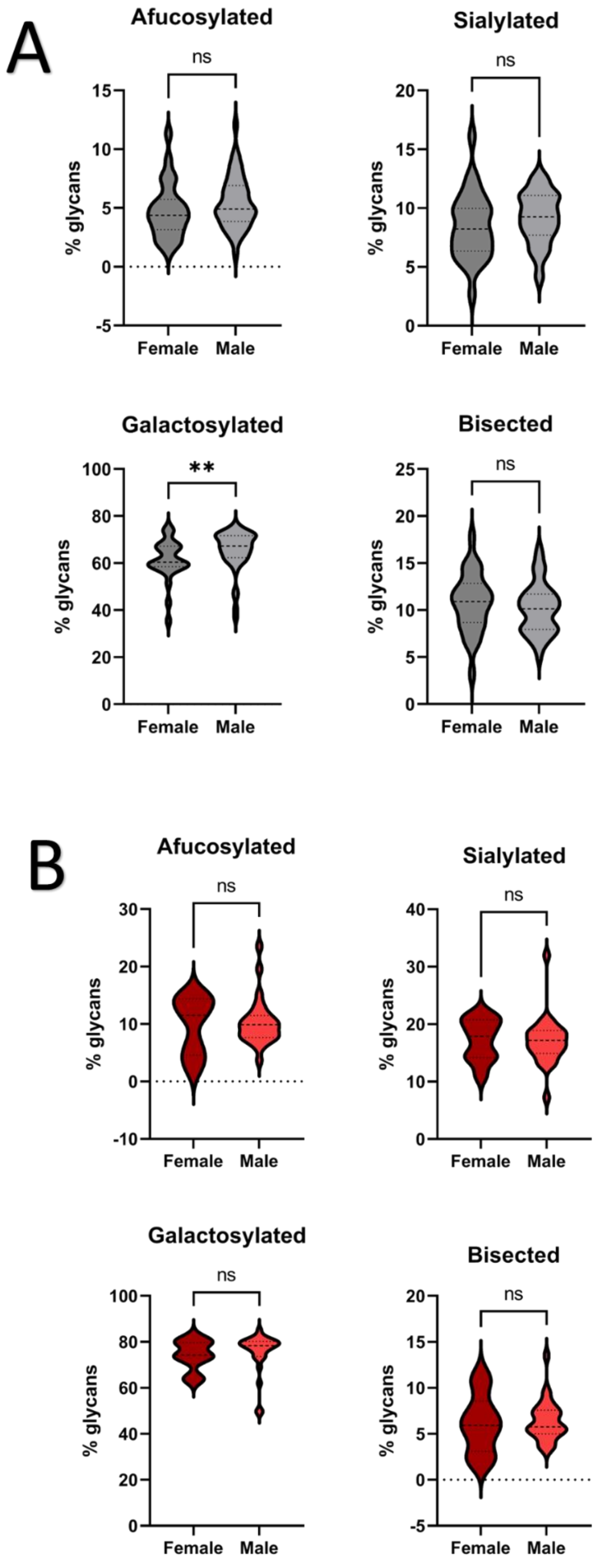


**Figure S8: Differences in IgG Fc glycans between sexes in adult COVID patients.** Differences in afucosylated, sialylated, galacosylated, or bisected **A)** bulk (gray) or **B)** anti-spike (red) IgG Fc glycan abundances between male and female adult COVID patients with moderate and severe COVID. Mann-Whitney test (* $p<0.05$, ** $p<0.01$, *** $p<0.001$, **** $p < 0.0001$)

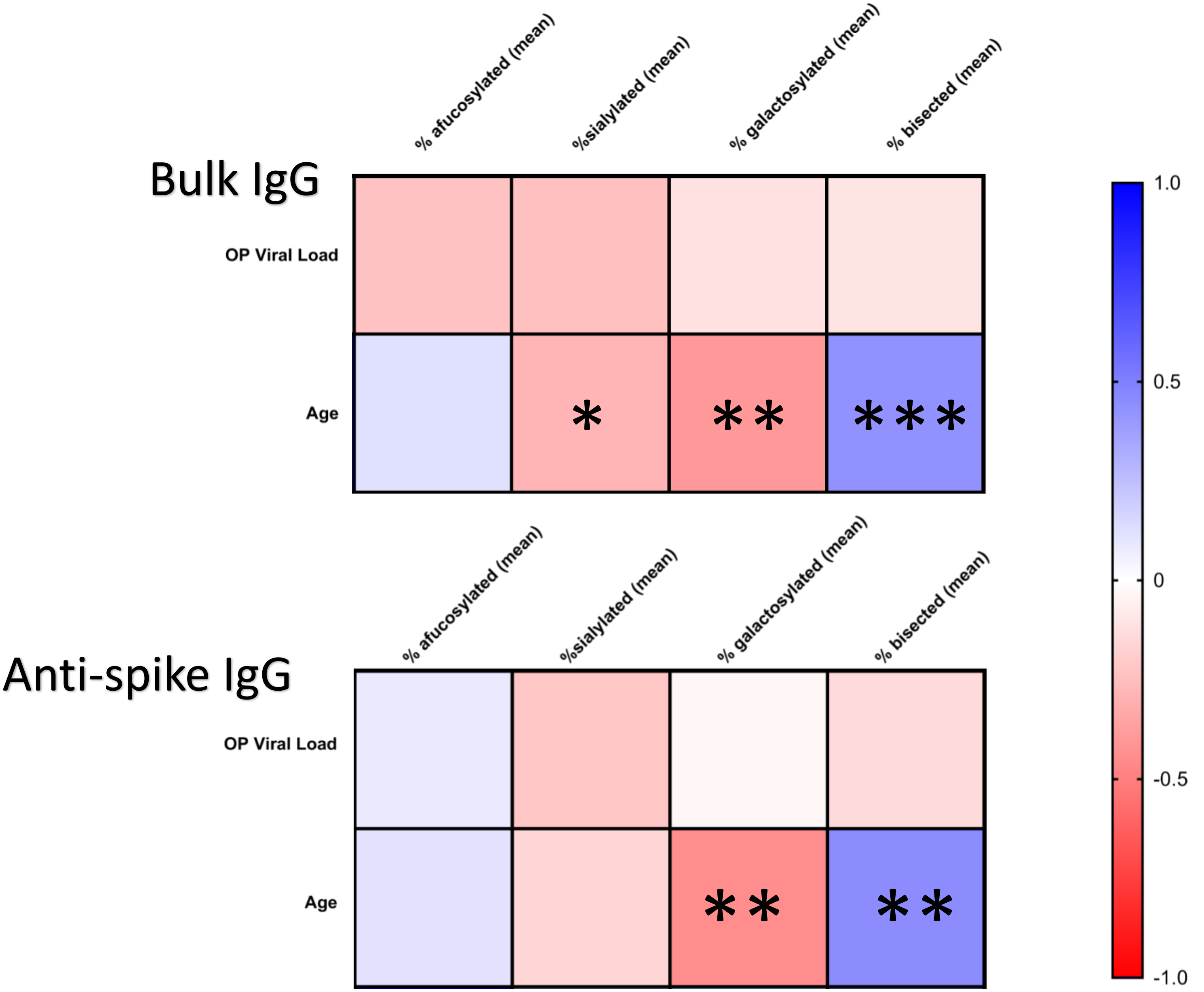


**Figure S9: Correlations observed between IgG Fc glycan abundances and age in adult COVID patients.** Spearman correlation coefficients were estimated between bulk/anti-spike IgG Fc glycan abundances, age, and oropharyngeal (OP) viral load in adult COVID patients. Gradient from blue to red indicates strength of positive or inverse correlation, respectively. Significant relationships are indicated by asterisks (* p<0.05, ** p<0.01, *** p<0.001).

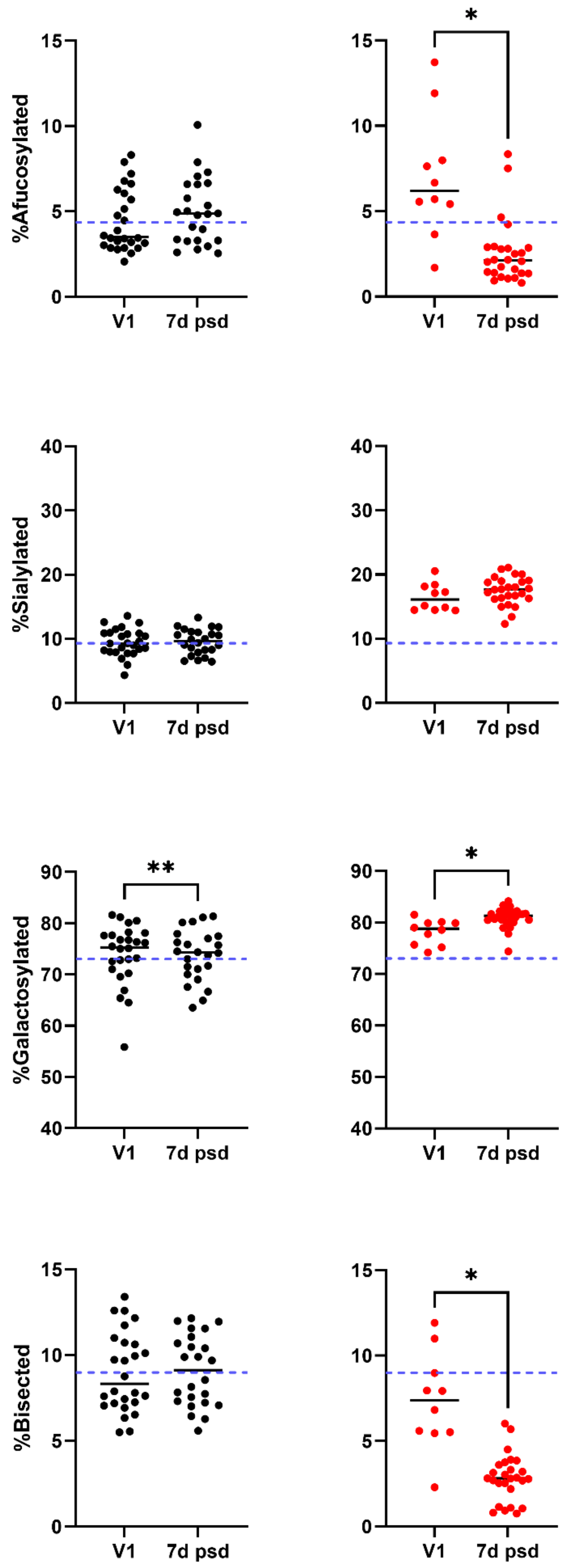


**Figure S10: Changes in IgG Fc-glycans after mRNA COVID vaccination.** Differences in afucosylated, sialylated, galacosylated, or bisected bulk (black) or anti-spike (red) IgG Fc glycan abundances between baseline (V1, before first vaccine dose) and 7 days post second vaccine dose (7d psd) in paired serum samples of adults who received an mRNA COVID vaccine. Wilcoxon matched-pairs signed rank test (* p<0.05, ** p<0.01, *** p<0.001, **** p < 0.0001)
